# Exposomics for childhood asthma

**DOI:** 10.64898/2026.03.02.26347385

**Authors:** Geoffrey L. Winsor, Justin Cook, Karlie Edwards, Erin E. Gill, Charisse Petersen, Karar Al-Mamaar, Amirthagowri Ambalavanan, Spencer Ames, Sonia S. Anand, Marie-Claire Arietta, Rilwan Azeez, Allan B. Becker, Lars Bode, Elissa Brookes, Darlene L.Y. Dai, Ruixue Dai, Bassel Dawod, Judah Denburg, Russel J. DeSouza, Dany Doiron, Qingling Duan, Thomas Eiwegger, Lauren Erdman, Kelsey Fehr, Catherine J. Field, Justin Foong, Emma Garlock, Anna Goldenberg, Emma Griffiths, Stan He, Michelle Helm, Perry Hystad, Meaghan J. Jones, Adrienne Kwan, Luis Ledesma Vega, Brenna Lee, Bryan Lee, Diana L. Lefebvre, Maxwell W. Libbrecht, Larisa Lotoski, Kelly McNagny, Luisa Mercado, Garthika Navaranjan, Krista M. Pace, Jaclyn Parks, David M. Patrick, Martin Pham, Yu Chen Qian, Myrtha E. Reyna, Hind Sbihi, James A. Scott, Michael Surette, Kristina Szabo, Tim K. Takaro, Piush J. Mandhane, Kozeta Miliku, Paul M. O’Byrne, Elinor Simons, Jeffrey R. Brook, Michael Brudno, Michael S. Kobor, Anita L. Kozyrskyj, Wendy Y. W. Lou, Theo J. Moraes, Malcolm R. Sears, Meghan B. Azad, Stuart E. Turvey, Padmaja Subbarao, Fiona S.L. Brinkman

**Affiliations:** Simon Fraser University; University of British Columbia; University of Manitoba; University of Cincinnati; McMaster University; BC Centre for Disease Control; University of Montreal; The Hospital for Sick Children, University of Toronto; University of California, San Diego; Queen's University; University of Toronto; Oregon State University; University of Calgary; University of Alberta

## Abstract

Identification of early interventions to reduce/eliminate asthma - the most common chronic disease among children - could significantly reduce burden on the healthcare system. Large-scale asthma Exposome-Wide Association Studies (ExWAS) could identify potential interventions, however integration of diverse data is required to address association confounders. The CHILD Cohort Study has followed 3,454 healthy Canadian children and their families from early pregnancy, collecting exceptionally diverse data including 27,006 variables from participant questionnaires, clinical data, household and neighbourhood-level exposures, and sample-derived chemical analytic/omic datasets. Here, we report integration of these datasets into the CHILDdb database platform, and use these data to perform ExWAS and machine learning analyses, identifying and further characterizing associations between childhood asthma and 2,954 diverse early exposures (pregnancy-age 5). Significant asthma associations include antibiotic use, human milk components, DEHP-phthalate, and mother’s prenatal cleaning product/disinfectant exposure. Subsequent analysis revealed epigenetic changes in the cord blood at birth, after prenatal cleaner exposure, and different microbiome and/or inflammatory cytokine changes associated with different asthma-associated exposures in the child. Collective results support asthma as a heterogeneous condition involving multiple etiologies, with associated endotypes, including significant prenatal exposures with potential transgenerational effects, and suggest targets for early interventions.

**Credit Roles:** 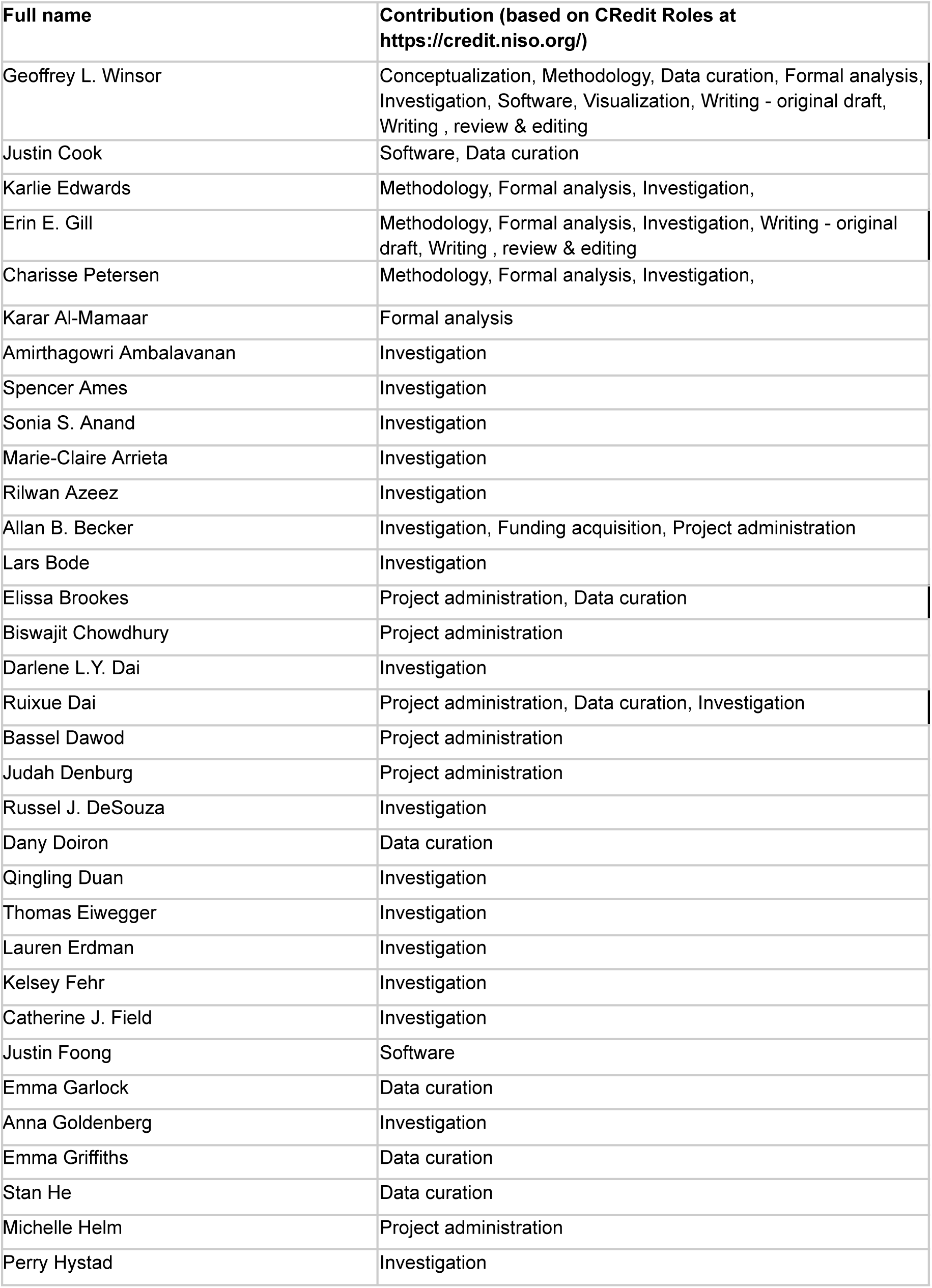

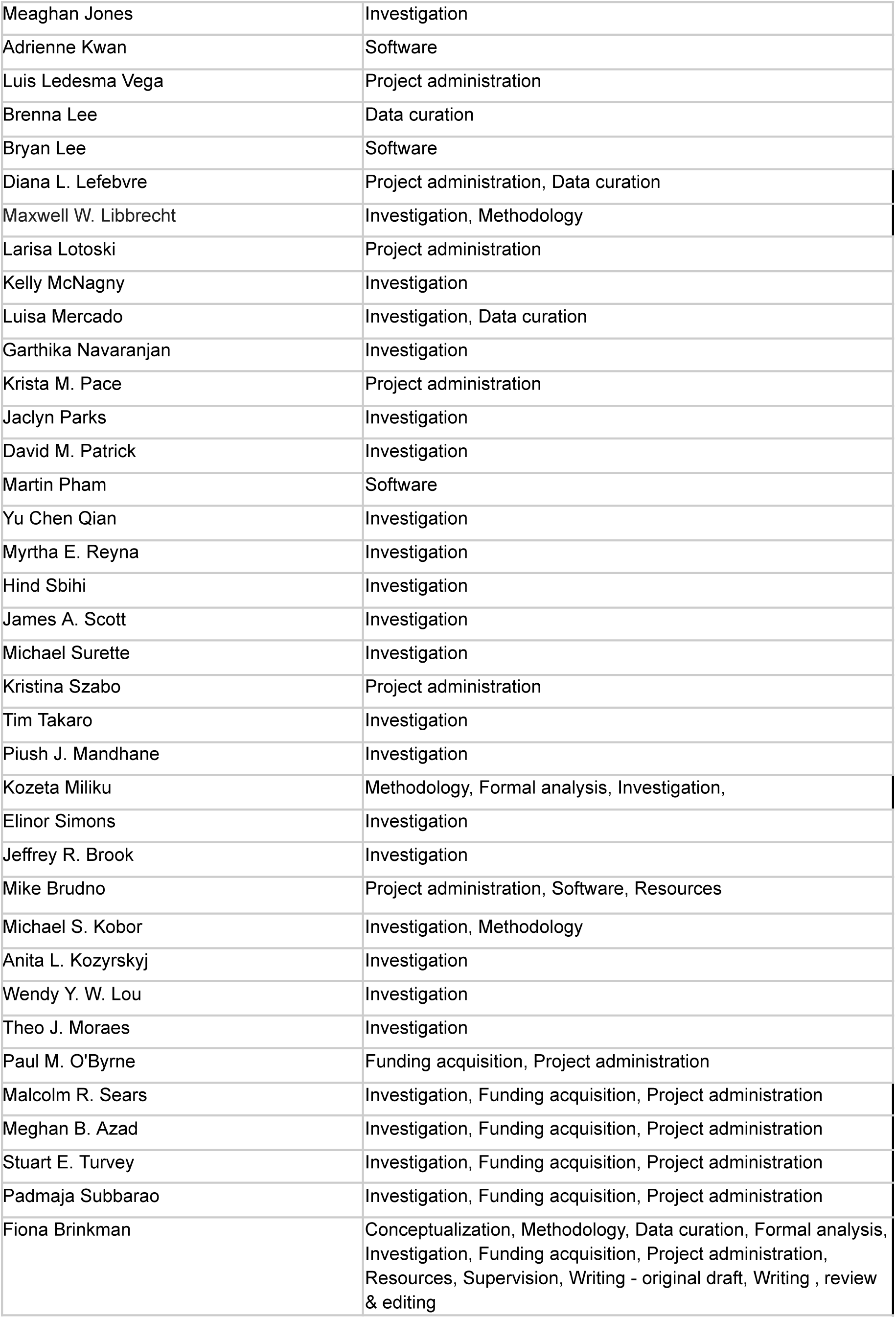

## Introduction

Non-communicable diseases rooted in early childhood often persist into later life^1–4^, emphasizing the need to identify early preventative interventions^5,6^. Asthma, the most common chronic disease in children, is a significant burden on the healthcare system and can persist into adulthood, with associated declining pulmonary function, and heightened risk for comorbidities ^7–12^. Asthma exhibits heterogeneity in phenotypes^10,11^, with diet, antibiotic use, air quality including smoking, and other environmental and lifestyle factors encountered during critical developmental windows appearing to significantly influence asthma risk^13–20^. Emphasis has been placed on the study of early postnatal exposures, however some exposures during pregnancy, including maternal stress, medications, diet, and tobacco smoke, have also been associated with increased asthma risk in a child^21–23^. Additionally, family asthma history and genetic variants influence susceptibility^24^.

Large-scale exposome studies, or exposomics (collective study of non-genetic factors impacting health or disease), have identified important asthma associations, including a French NutriNet-Santé cohort study of adults, an INMA cohort study focused only on phthalates, and a New Bedford study focused only on chemicals. However, these studies have been limited in terms of the variety of exposures examined, the diversity of the participants, or the size of the cohort^25–27^. Exposomics must also move toward identification of more causal factors of disease – more adequately factoring in the interrelationships between different disease-associated exposures with each other. This requires harmonized integration of very diverse, longitudinally-collected exposure measurements for a suitably sized, relatively genetically diverse, population-based cohort^23,27–31^.

The CHILD Cohort Study (CHILD) represents a unique opportunity to more integratively characterize a wider range of exposures and support more integrative exposomics^30^. CHILD is a large and long running Canadian prospective longitudinal birth cohort, tracking exceptionally diverse data types per participant, with over 27,000 data points on average per participant for 3,454 children and their families. The families generally reflect the demographics of Canadians (urban vs rural, and ethnic diversity, though with a moderate bias towards higher SES common in many cohorts)^32^. CHILD represents more ethnic diversity, or larger cohort size, than cohorts used in asthma exposome studies to date^33^. CHILD’s recruitment of healthy participants at birth, and collection of data at very early critical development milestones is providing insights into the roles that environmental exposures, microbiomes, genetics, and epigenetics play in the development of asthma, allergy and other childhood diseases^34–38^ ^31,39^.

We now report development of an integrative data platform called CHILDdb that combines harmonized data from the four CHILD sites, spanning the 18th week of pregnancy to 8 years of age, with ongoing longitudinal data integration. We use data from CHILDdb to perform an asthma Exposome-Wide Association Study (ExWAS), and machine learning analyses, using 2,954 environmental and lifestyle variables, including products used in the home, human milk components and other nutritional information, chemical analytes, and neighbourhood-level environmental exposures. Asthma-associated environmental factors were integratively studied with gut microbiome, epigenetic, and inflammatory marker datasets, to further explore putative disease mechanisms and support hypothesis generation for causation testing.

The collective results highlight the importance of employing a more unbiased approach that integrates diverse prenatal and postnatal environmental factors, identifying early exposures and hypotheses for mechanisms for childhood asthma development - paving the way for timely early, appropriate interventions that can reduce healthcare burden due to asthma.

## Results and Discussion

### CHILDdb integrates diverse environmental, health, omics, and phenotypic datasets

The CHILDdb.ca database, plus its web platform for viewing CHILD variables and making data requests, is one of the most diverse data platforms of its kind to date. CHILDdb currently integrates 27,006 variables per participant, from 242 diverse datasets, harmonized across four study centres across Canada (Figure 1; childdb.ca). Data has been collected primarily at 12 time points from about the 18th week of pregnancy to age 8 of the child, including a COVID-19-add-on study^40^, while integration of age 13 data and collection of age 16 data is ongoing. Data includes validated questionnaire responses, technician-based home assessments, clinical assessments by asthma/allergy specialists, geospatial data from CANUE: the Canadian Urban Environmental Health Research Consortium^41^ linked by postal codes and then anonymized (including socioeconomic and neighbourhood environmental data), omics data (Supplementary figure 1; including microbiome, epigenome and genetic data), ontologies and other controlled vocabularies to enable organized hierarchical analysis of data (e.g. ChEBI for medications) and sample-derived chemical analytic data (urine, stool, serum, breastmilk, house dust). As a leading prospective, longitudinal birth cohort, CHILD has increased understanding of the early-life exposures impacting asthma and other common allergic diseases^17,42–44^ (https://childcohort.ca/publications/).

**Fig.1.**
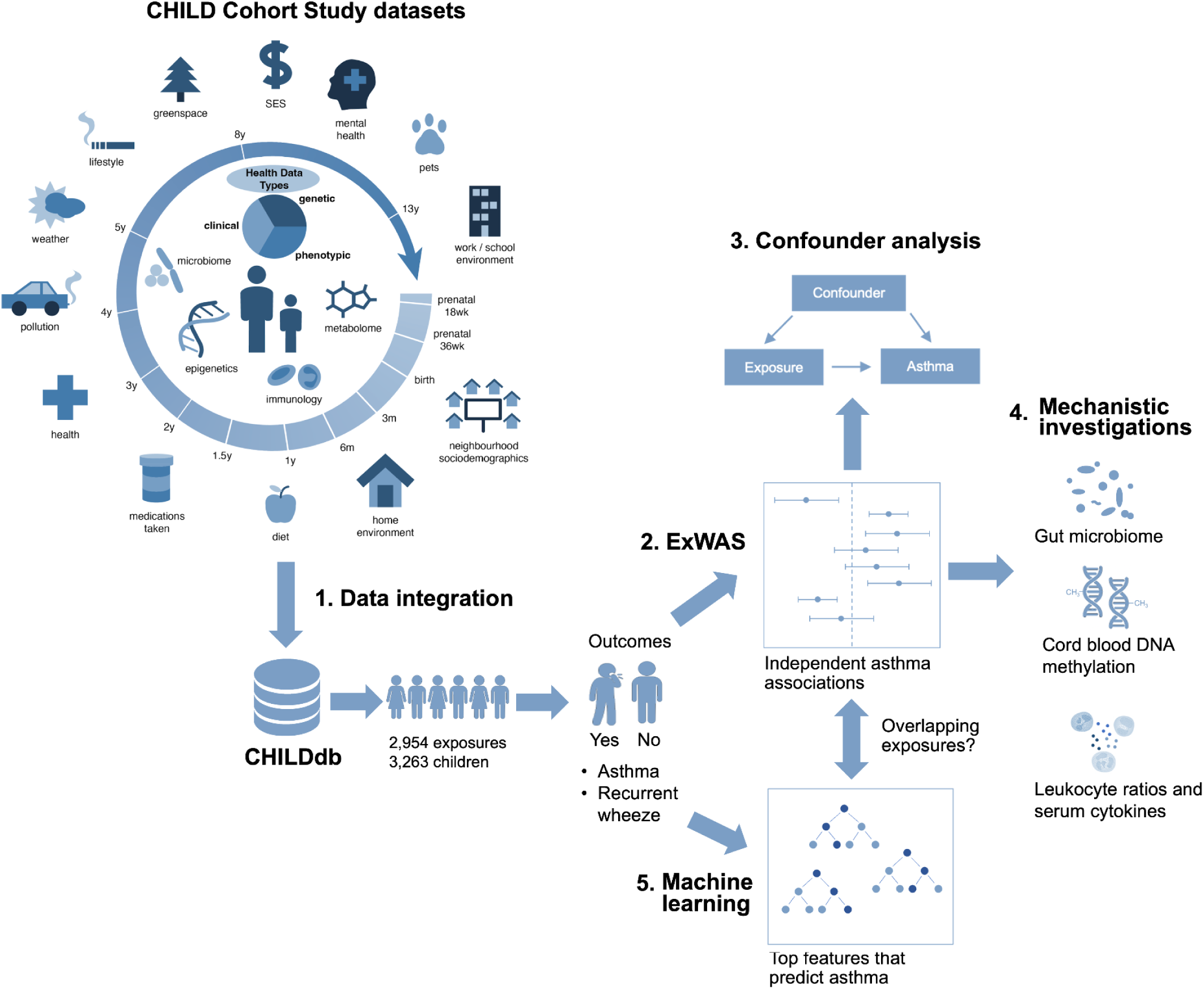
CHILD Cohort Study data integration enables characterization of associations with the early-life exposome. Data collected at time points ranging from enrollment during pregnancy, to the 8th year visit were integrated into CHILDdb (13-year visit data has been collected and is currently undergoing cleaning). Sample-derived data (inside the ring) includes gut, nasal, and breast milk microbiome, epigenetic, metabolomic, and immunological data, along with clinical test results, participant phenotypes and genetic variant data. In addition it contains diverse questionnaire-derived datasets and external-derived socioeconomic and environmental datasets linked to securely stored, anonymized participant postal codes. After integration in CHILDdb, a dataset of 2,954 exposures (most of the exposures encountered between pregnancy and 3 months of age) was used for an exposome-wide association study (ExWAS) analysis to identify independent associations with asthma at the age of 3 years (n=2,544), 5 years (n=2,399), and 8 years (n=2,293) or recurrent wheeze at the age of 1 year (n=3,144), 3 years (n=2,856), and 5 years (n=2,719). Significant exposures underwent more stringent analysis with additional covariates added in order to further confirm the associations. These exposures were then subjected to additional mechanistic investigation to provide evidence for putative mechanisms of action. In order to provide more support for exposures identified by ExWAS, additional machine learning approaches were used to predict asthma from a dataset of primary exposures (hypothetical initial exposures) and features that were most important for the model were identified.

Compared to some other well-established child cohorts worldwide^45–51^, CHILD captures greater racial and ethnic diversity (approximately 28% of participants were identified as belonging to racialized groups), has a larger cohort size, or more early-life measurements, while reflecting Canadian demographics^38^. The CHILDdb.ca portal facilitates requests for its complex individual-level data, including an elasticsearch engine (https://www.elastic.co/elasticsearch) to support advanced queries (with criteria to prevent participant identification), data visualizations which enable equitable studies with statistical power, functionality enabling full administrative and executive review of data requests from researchers, and support for collaboration between data requesters/analyzers and data generators. Source code is freely available under an open source licence.

### An Exposome-Wide Association Study of prenatal and postnatal exposures supports a multifactorial etiology for childhood asthma

Capitalizing on CHILDdb, an Exposome-Wide Association Study (ExWAS) using logistic regression assayed 2,954 exposure variables, from 25 different categories of exposures in CHILDdb (Supplementary table 1), for associations with nine specialist clinician-diagnosed asthma and recurrent wheeze endpoints in children aged 1 to 8 years (Figure 1; Supplementary table 2). After adjusting for covariates biological sex^52^, study site and season of birth^53^, 126 distinct exposure variables were identified as significant over the nine asthma/wheeze endpoints examined after correcting for multiple testing (Supplementary file S1A). In a separate Spearman rank correlation analysis, the covariates season of birth and study site were further analyzed, to confirm their associations, and significantly correlated (absolute value Spearman rho >= 0.3) with precipitation, temperature, and previously reported risk factors for developing/exacerbating asthma (e.g. nitrogen dioxide, PM2.5, ozone^54–56^), consistent with the site diversity across Canada (Supplementary file S1B).

A second, more stringent ExWAS analysis was then performed on these significant exposures, adjusting for previously-established covariates comprising family asthma history^57^, antibiotic use^17^, presence of older siblings^16^, household income^58^, breastfeeding^59^, birth mode^60^, any prenatal smoking in the household^61^, study site, season of birth^53^ and biological sex^52^, to identify any novel associations. After correction for multiple testing, most (112 of 126) exposures remained significantly associated collectively with the nine asthma and/or recurrent wheeze endpoints (Supplementary file S2).

As expected, established risk factors for asthma (male biological sex, family history of asthma) and a more recently-appreciated risk factor (antibiotic use) were positively associated with specialist clinician-diagnosed ’definite’ asthma at age 5 (Figures 2a and 2b). Also, use of albuterol (salbutamol) or anti-allergenic drugs (medications for treatment of recurrent wheeze/asthma^62^ or allergies, respectively) were positively associated - consistent with indication bias (a form of “reverse causation” where, for example, albuterol is prescribed because of asthma symptoms rather than causing asthma). Similarly, certain solid foods (“yes/no” questions) including eggs, peanuts, nuts and dairy products consumed between 18 and 30 months of age were negatively associated with asthma, potentially reflecting exposure to common food allergens, since food allergy is associated with asthma development^63^. However, though reverse causation may be a factor, further study of the role of such foods is needed.

**Fig. 2.**
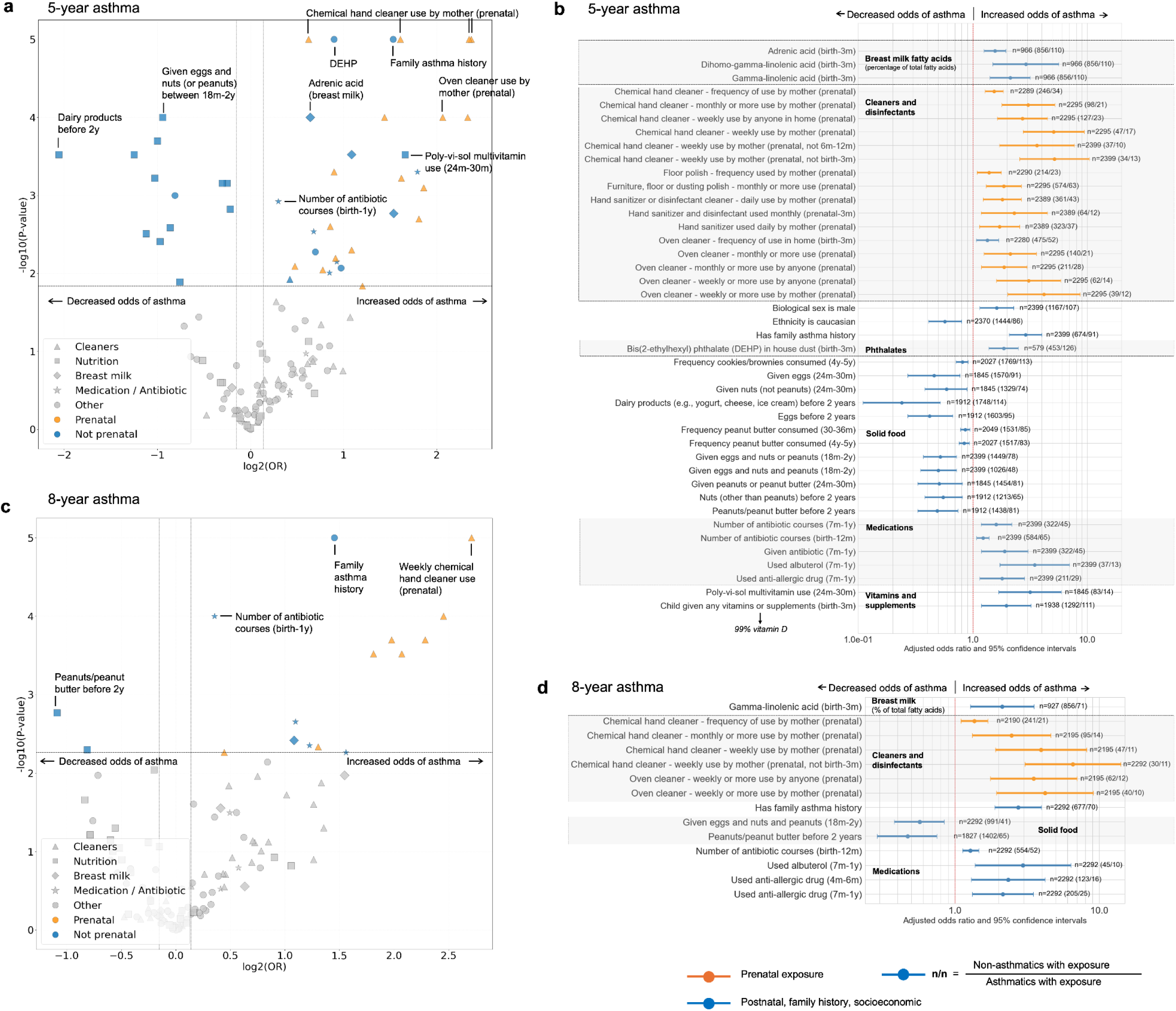
Exposures identified by ExWAS as associated with specialist clinician-diagnosed asthma at 5 and 8 years of age. **a.** and **c.** Volcano plots showing exposures that are significantly associated with 5- and 8-year year asthma, respectively, after a second more stringent ExWAS (results of the exploratory first round of ExWAS are found in Supplementary file S1A).Logistic regressions controlling for additional covariates were conducted on the 112 variables significantly associated with asthma or wheeze in the first round. Significant prenatal exposures are coloured orange while other significant exposures are blue. Shapes reflect the main categories of exposure. **b.** and **d.** Forest plots of the same exposures that were significantly-associated with 5- and 8-year year asthma, respectively, after the second round of ExWAS. Orange and blue horizontal plots are adjusted odds ratios and 95% confidence intervals for prenatal and postnatal exposures, respectively. Positive associations have odds ratios over 1 and negative associations are less than 1. Numbers to the right of each plot represent the total N for each individual logistic regression analysis followed in brackets by the number of non-asthmatic cases / asthmatic cases that were subjected to each exposure, respectively. All were significant after correcting for multiple testing (see methods). Major groups of exposures are labelled and coloured for additional clarity (Grey).

The ExWAS identified other exposures positively associated with 5-year asthma, that are not well characterized in the literature. These included the proportion of three omega-6 fatty acids in breast milk, DEHP (bis(2-ethylhexyl)) phthalate levels in house dust (found in certain plastics), at least one vitamin or supplement being given to the infant between birth and 3 months of age, and frequent maternal cleaning product use during the prenatal period (Figures 2a and 2b).

Strikingly, daily prenatal use of hand sanitizer (OR: 1.71, 95% CI: [1.14, 2.56]) and multiple different cleaning products were among the highest odds ratios. Cleaning products included prenatal weekly oven cleaner and prenatal weekly chemical hand cleaner (e.g. hand degreaser - chemicals used frequently in house cleaning and related occupations) - the latter having a significantly higher odds ratio than antibiotic use, birth to 12 months (or other measures of antibiotic use; OR: 3.12, 95%, 95% CI: [1.29, 7.56] for the cleaner, versus OR: 1.24, 95% CI: [1.05, 1.46] for antibiotic use; Wald test for difference in coefficients p <0.05). This frequent use, specifically maternally and prenatally, is correlated with the development of asthma, even if the mother stopped exposure after birth (as might occur if the exposure was occupational before maternity leave). Cleaning product use at multiple other postnatal timepoints between birth and 5 years of age was not significant, supporting the significance of such maternal prenatal cleaning product exposure and the development of asthma. Earlier wheeze and asthma endpoints were associated with more exposure variables and categories than at age 5. Recurrent wheeze at age 1 year was also positively associated with early postnatal cleaning product use, air fresheners, pre- and postnatal tobacco smoke, and respiratory infections (Supplementary figure 2). Notably, in contrast to the positive association with asthma at age 5 years, vitamin and/or supplement use was negatively associated with recurrent wheeze at the age of 1 (OR: 0.60, 95% CI: [0.43, 0.84]; Supplementary figure 2). Recurrent wheeze and asthma at age 3 were positively associated with frequent use of baby lotion (possible proxy for atopic dermatitis), hours spent per weekday in a vehicle (possible proxy for traffic pollution), and postnatal exposure to cleaning products/cleaning of homes 4 or more times per month (Supplementary figures 3 and 4). The increased number of associations at ages 1 to 3 is consistent with preschool wheezing being a more heterogeneous condition, which makes accurate 3-year asthma diagnoses challenging^64^. The results are also consistent with previously reported associations with wheeze and lung function at a younger age, including tobacco smoke exposure^61^, vehicle exhaust^65^ and frequent postnatal cleaning product use^66^. Conversely, asthma diagnoses at ages 5 and up are based on less heterogeneous clinical presentation that match the underlying pathology, while factors that contribute to earlier diagnoses (e.g. viral infections, males with smaller airways) have been observed to improve, or the diagnoses are relabelled over time^64,67^. At age 8 years, only twelve exposure variables were positively associated and two exposures negatively associated with asthma (Figures 2c and 2d), however, they all overlapped with exposures significant in the 5-year analysis. Again, weekly prenatal chemical hand cleaner use was associated with higher odds of 8-year asthma than infant antibiotic use from birth to 12 months (OR: 4.73, 95% CI: [ 2.11, 10.63] vs. OR: 1.32, 95% CI: [1.16, 1.51]; Wald p<0.05) - including mother’s use prenatally but not after birth. Collectively, there was a general lack of correlation between different “categories” of asthma-associated exposures, including those still found significant after controlling for each other: prenatal cleaning product use, phthalate exposure, antibiotic use, and breast milk components (Supplementary Results, Supplementary Figure 5). This is consistent with the paradigm that multiple causative factors and exposure pathways can increase the risk of asthma, and may converge on the same or similar eventual clinical phenotype, despite potentially different underlying biological/inflammatory processes.

### Frequent use of cleaning products and disinfectants during pregnancy is associated with downstream epigenetic changes in cord blood and enriched pathways characterized by neutrophilic asthma cytokine profiles later in the child

Despite assessment of cleaning product exposures from early pregnancy through 5 years of age, most significant associations with asthma at age 5 were observed during the prenatal period. These associations persisted after factoring in postnatal exposure to the same cleaning products (Supplementary results, Supplementary figure 6).

Given the prenatal timing of these exposures, DNA methylation profiles from cord blood were examined in an epigenome-wide association study where significant associations were observed between prenatal use of hand sanitizer, chemical hand cleaner, oven cleaner, and other disinfectants (Figure 3, Supplementary file S4). The magnitude of change of these associations was similar to those previously described following prenatal smoke exposure^68,69^, although smoke exposure did not correlate with any cleaning product use and different genes have been identified as associated with smoke exposure^69,70^.

**Fig. 3.**
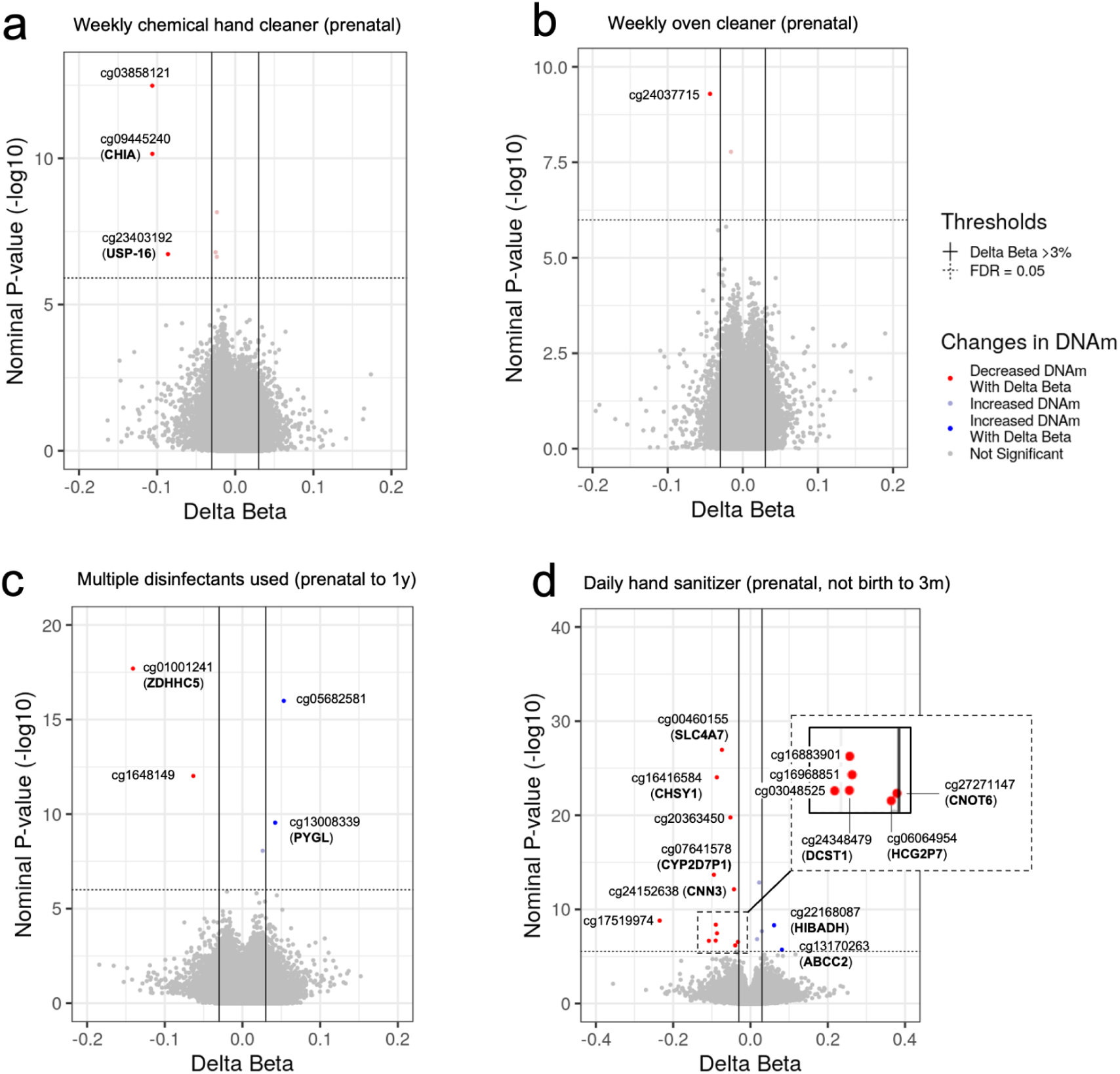
Use of cleaning/disinfecting products corresponds with differential methylation at CpG sites within the genome. **a**. Weekly chemical hand cleaner used by mother (prenatal). **b.** Weekly oven cleaner used by mother (prenatal). **c.** Use of multiple disinfectants on a monthly or more frequent basis by mother during pregnancy and in home up to 1 year of age (defined by affirmative responses to having used hand sanitizer, disinfectant cleaner for general house cleaning, and disinfectant cleaner for the bedroom). **d**. Daily hand sanitizer used by mother (prenatal) and monthly or less from birth to 3 months. Red dots are CpG sites in DNA with significantly decreased methylation while blue dots are CpG sites with significantly increased methylation. CpG sites are labelled with the gene they are located in, if applicable.

Most notably, weekly or more chemical hand cleaner use by the mother during pregnancy, which was associated with increased odds of having asthma at 3, 5 and 8 years, was also significantly associated with reduced methylation at three CpG sites (Figure 3a), including cg09445240 in the CHIA gene, which encodes acidic mammalian chitinase, that co-localizes at sites of Th2-mediated inflammation and stimulates chemokine production by pulmonary epithelial cells^71^. Also significant was cg23403192 in the USP16 gene, encoding a ubiquitin specific peptidase involved in activation and proliferation of T cells.

A subcohort of mothers who used hand sanitizer daily during pregnancy, but not at all between birth and 3 months (suggesting occupational use), had multiple significant changes in DNA methylation (Figure 3d), including decreased methylation at a CpG site within CHSY1, a gene associated with increasing the levels of macrophages and neutrophils in mice^72^. There were also associations with decreased methylation within DCST1, which plays a role in negative regulation of type 1 interferon signaling^73^, and bicarbonate transporter SLC4A7, which regulates macrophage phagosome acidification^74^. To our knowledge, this is the first study reporting epigenetic changes in cord blood associated with prenatal hand sanitizer and other cleaning product use.

There was no evidence for mother’s prenatal cleaning product use being associated with any subsequent change in gut microbiome alpha diversity in their children at 3 months and 1 year. Prenatal oven cleaner (weekly or more use) and hand sanitizer (daily use) by the mother were associated with changes in 3-month and 1-year abundance of some taxa, however the magnitude of change and number of taxa were similar to that observed for antibiotic use exposure (birth to 3 months and birth to 1 year, respectively), but modest compared to changes associated with duration of breastfeeding, birth mode, and having an older sibling (Supplementary figure 7).

However, for mothers who used hand sanitizer daily during pregnancy, their children had on average higher levels of serum adenosine deaminase at age 1 (a measure of inflammation; Supplementary figures 8a-8b). Analysis of the child’s serum, using a Olink protein marker panel, coupled with Gene Set Enrichment Analysis (GSEA) identified cytokine levels increased at 5 years that were enriched for pathways including maturation of monocyte-derived dendritic cells (a cell type that has been implicated in atopic asthma pathogenesis^75^). Gene Ontology (GO) biological processes enriched in the GSEA included regulation of cell migration, chemotaxis, and response to interleukin-1, and the “IL-10-downregulated extracellular proteins” portion of the IL-10 signalling pathway – consistent with previous studies showing that reduced IL-10 signalling is associated with increased disease severity in asthmatics^76^ (Supplementary figure 8c).

The leading edge cytokines driving these enriched pathways (Table 1) have been previously attributed to a Type 2 low (T2-low) immune response, characteristic of neutrophilic asthma and driven by CD4+ Th17 T cells and interleukin-6, though a more severe CD4+ Th1-mediated variant involving more IFN-γ has also been described^77,78^ (Supplementary table 3). For a subset of children in this cohort, our findings putatively suggest an association with neutrophilic asthma. This endotype has been linked to oxidizing chemical exposures and is notable for its resistance to corticosteroid-based treatment^79–83^.

**Table 1.**
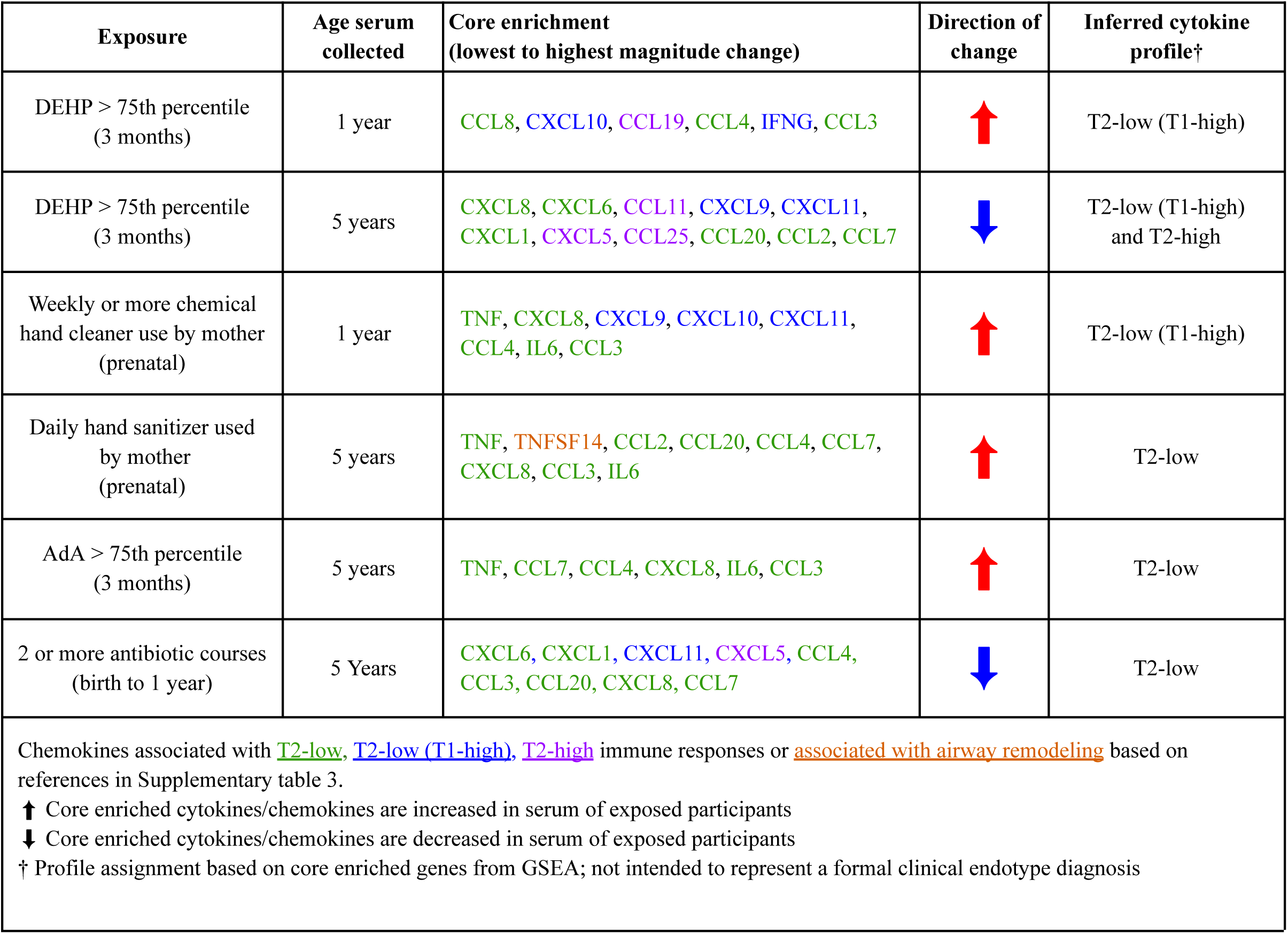
Characteristics of core enriched (leading edge) biomarkers of pathways from 1- and 5-year serum of children exposed to: bis(2-ethylhexyl) phthalate (DEHP) levels in house dust > 75th percentile, weekly or more chemical hand cleaner (e.g. hand degreaser) use by mother (prenatal), daily hand sanitizer use by mother (prenatal), breast milk adrenic acid (AdA) levels > 75th percentile, and 2 or more courses of systemic antibiotics taken between birth and 1 year. Associations between each individual chemokine and immune profiles (Th2-high, Th2-low, Th2-low/Th1-high, or association with airway remodelling) were curated from scientific literature and a putative overall endotype assigned.

| Exposure | Age serum collected | Core enrichment (lowest to highest magnitude change) | Direction of change | Inferred cytokine profile† |
| --- | --- | --- | --- | --- |
| DEHP > 75th percentile (3 months) | 1 year | CCL8, CXCL10, CCL19, CCL4, IFNG, CCL3 | ↑ | T2-low (T1-high) |
| DEHP > 75th percentile (3 months) | 5 years | CXCL8, CXCL6, CCL11, CXCL9, CXCL11, CXCL1, CXCL5, CCL25, CCL20, CCL2, CCL7 | ↓ | T2-low (T1-high) and T2-high |
| Weekly or more chemical hand cleaner use by mother (prenatal) | 1 year | TNF, CXCL8, CXCL9, CXCL10, CXCL11, CCL4, IL6, CCL3 | ↑ | T2-low (T1-high) |
| Daily hand sanitizer used by mother (prenatal) | 5 years | TNF, TNFSF14, CCL2, CCL20, CCL4, CCL7, CXCL8, CCL3, IL6 | ↑ | T2-low |
| AdA > 75th percentile (3 months) | 5 years | TNF, CCL7, CCL4, CXCL8, IL6, CCL3 | ↑ | T2-low |
| 2 or more antibiotic courses (birth to 1 year) | 5 Years | CXCL6, CXCL1, CXCL11, CXCL5, CCL4, CCL3, CCL20, CXCL8, CCL7 | ↓ | T2-low |
| Chemokines associated with T2-low, T2-low (T1-high), T2-high immune responses or associated with airway remodeling based on references in Supplementary table 3.<br>↑ Core enriched cytokines/chemokines are increased in serum of exposed participants<br>↓ Core enriched cytokines/chemokines are decreased in serum of exposed participants<br>† Profile assignment based on core enriched genes from GSEA; not intended to represent a formal clinical endotype diagnosis |  |  |  |  |

Children of mothers who had used chemical hand cleaner weekly or more during pregnancy had a 1-year serum cytokine profile that was enriched for the toll-like receptor signalling pathway (KEGG:04620, WikiPathways:WP75), based on a core enrichment set containing inflammatory cytokines that are all annotated as being released downstream of either the MyD88-dependent TLR pathway or the JAK/STAT component of the MyD88-independent TLR pathway. These biomarkers are associated with a T2-low (Th1-high subset) profile (Table 1, Supplementary figure 9a), including CXCL9, CXCL10, and CXCL11, which are strong indicators of an IFN-γ response^84^. In contrast to hand sanitizer and chemical hand cleaner, weekly or more prenatal oven cleaner use was associated with higher eosinophil-to-lymphocyte (β=0.264, 95% CI: [0.013, 0.514]) and eosinophil-to-neutrophil ((β=0.344, 95% CI: [0.0.063, 0.625]) ratios at age 1 year (linked in other studies to eosinophilic asthma^85–87^) but not associated with enrichment of any inflammatory pathways (Supplementary figure 9b-9e).

The associations between child asthma and mother’s use of prenatal cleaning products is supported by evidence from more limited studies in other cohorts: Using a Total Chemical Burden (TCB) approach, the ALSPAC cohort found a positive association between prenatal cleaner use and development of wheeze at 42 months, but did not rule out any contribution from postnatal exposure^88^. A later study by ALSPAC used an index similar to TCB called Composite Household Chemical Exposure (CHCE) score to show that non-transient wheezing up to 7 years of age is associated with frequent use of chemicals in the home but did not determine if this was a proxy measure for cleaner use during the entire postnatal period, nor identify individual products^89^. Limited additional studies support associations between occupational use of more general ‘medical disinfectants’ during pregnancy and self-reported asthma or wheeze at early ages^90,91^.

While most cleaning product associations with childhood asthma and wheeze in our study were prenatal in origin, CHILD researchers in another study used a Frequency of Use Score (FUS) to demonstrate that use of multiple cleaning products during infancy (birth to 3 months) increased the odds of recurrent wheeze or asthma at 3 years, but did not examine prenatal exposure nor examine later time points^66^. Corroborating results from this study, we found that infants from homes that had 4 or more general cleanings per month (0-3m) did have increased odds of developing asthma and recurrent wheeze at 3 years of age (Supplementary figures 3, 4) and this was associated with increased serum IL-4, IL18R1, and Flt3L (Supplementary figures 10a, 10b) and enrichment of GO Biological Processes including “Response to Cytokines” and “Defense Response” at 5 years (Supplementary figure 10d).

Collectively, this work more comprehensively characterises associations between exposure to prenatal cleaners and subsequent childhood asthma or recurrent wheeze (ages 1 to 8 years), while factoring in postnatal cleaning product use and a diverse dataset of other exposures. This is also the first study to identify and describe epigenetic changes associated with this exposure that are highly significant and warrant further study. Notably, frequency of home cleaning was not correlated with hand sanitizer, oven cleaner or chemical hand cleaner use, consistent with these latter frequent cleaners’ use in an occupational context (ie. house cleaners; example: Supplementary table 4). Similarly, frequent prenatal hand sanitizer use was associated with working in healthcare occupations (47% used it daily; data not shown) – a different group of mothers than those frequently using other cleaners: Venn diagrams of participants with asthma-associated frequent hand sanitizer use, chemical hand cleaner use, and antibiotic exposure do not commonly overlap (Supplementary figure 11).

It should be emphasized that if these cleaning product exposures are playing a causative role for asthma, the specific components are unknown. However, bleach or related cleaners are known to cause inflammation in respiratory epithelium^92^ and direct DNA base chemical changes at CpG sites, along with other impacts^93^. Hand sanitizer fragrances can contain phthalates, and phthalates may leach from plastic containers^94^.

### DEHP levels are positively associated with inflammation and childhood asthma

Among a subset of participants (n = 549; 118 with asthma, 431 without), increasing concentrations of bis(2-ethylhexyl) phthalate (DEHP) in house dust collected at a 3-month home visit were positively associated with the development of asthma at age 5 (OR: 1.86, 95% CI: [1.38, 2.5]; Figure 2b), even after factoring in other associated exposures. Similar to the prenatal oven cleaner findings, dust DEHP concentrations above the 75th percentile were associated with significantly higher eosinophil-lymphocyte (β=0.27, 95% CI: [0.08, 0.46]) and eosinophil-neutrophil (β=0.27, 95% CI: [0.04, 0.49]) ratios in age 1 whole blood (Supplementary Figures 12a,12b). GSEA identified upregulated biomarkers of GO biological processes including cation transport and metal ion transport (Supplementary figure 12c), and a T2-low (T1-high) cytokine profile (Table 1), in age 1 year serum. In 5-year serum, pathways related to chemokine and chemokine receptor binding^95^, genes upregulated by TNF, and granulocyte chemotaxis were negatively enriched (normalized enrichment scores < 0; Supplementary Fig. 12d). Unlike the other exposures undergoing GSEA, the enriched pathways for high dust DEHP were driven by downregulated core enrichment genes, specifically proinflammatory cytokines and chemokines shared across all three inferred immune profiles (T2-low, T2-low/T1-high, and T2-high; Table 1). More study is needed, however, DEHP and its metabolite MEHP have been previously implicated in the inhibition of chemotaxis,^96,97^ and DEHP has been linked to eliciting a type 2 inflammatory response in human macrophages^98^.

There were no associations between dust DEHP levels and any infant gut microbiome changes, nor significant epigenetic changes in cord blood DNA (data not shown). DEHP was positively correlated with monthly nitrogen dioxide (NO_2_) levels mapped to the participant family postal code at 9 months prior to birth and 12 months post birth (Supplementary figure 5a). Ambient NO_2_ levels between the prenatal to first year postnatal period have been linked to childhood asthma^93^, and increasing concentrations are correlated with higher levels of DEHP in house dust^100^. DEHP’s association with asthma though is maintained when adjusting for a large set of asthma-associated variables (Supplementary figure 5b) including pollution/smoke exposure. Phthalates have been associated with increased lung inflammation and changes in inflammatory cell numbers^101^. This association mirrors the results of a more limited earlier study with reduced confounders performed with the CHILD cohort^102^. Other cohort analyses of phthalates measured in urine, as well as mouse studies of phthalate exposure, support phthalate associations with asthma/impacts on asthma-associated genes, and even suggest a role for epigenetic changes with potential transgenerational effects^99–101^. Collectively, the inflammatory pathway associations and other findings for this exposure are consistent with DEHP acting through a distinct mechanism from the prenatal cleaning product exposures in at least a subset of the children studied.

### Vitamin D supplementation is associated with decreased gut microbiome alpha diversity

72% of infants (1403/1938) assessed for asthma in the 5-year ExWAS analysis were given at least one vitamin or supplement between birth and 3 months of age. This included vitamin D for 99% of the 1403 infants receiving vitamins or supplements, and is a standard of care recommended by Health Canada for breastfed babies^106^. The 5-year ExWAS showed that this supplementation was positively associated with asthma at age 5 (OR: 1.96, 95% CI: [1.19, 3.22]). However, after adjusting for an expanded set of covariates, including other asthma-associated exposures that correlated with vitamin D (including DEHP; Supplementary Figure 5a), the association was no longer observed (OR: 1.28, 95% CI: [0.87, 1.87]; Supplementary figure 5b, with similar analyses done for other asthma-associated exposures).

Supplementation between birth and 3 months was negatively associated with recurrent wheeze at age 1 (OR: 0.60, 95% CI: [0.43, 0.84]; Supplementary figure 2).

Most strikingly, vitamin D use was also associated with significantly reduced alpha diversity for the infant microbiomes at 3 months of age (Supplementary figure 13a), particularly in exclusively breastfed infants (e.g. Chao1, β=-8.37, 95% CI: [-10.72, -6.02]; Shannon, OR:-0.26, 95% CI: [-0.38, -0.15], Inverse Simpson, OR: -1.49, 95% CI:[-2.04, -0.93]) after stratification by breastfeeding status (Supplementary figure 13b).

In this study, 98% of vitamin D taken was brand name Enfamil D-Vi-Sol drops containing 400 IU/ml vitamin D3 and non-medicinal ingredients glycerin (carrier) and polysorbate 80 (emulsifier)^107^. The potential impact of Vitamin D supplementation needs further study, including its solvent carrier, considering that carrier type (oil versus water-based) has been suggested to impact risk of asthma^108^.

### Sex-specific association between proportion of adrenic acid in human milk and later childhood asthma

A protective association was observed between breastfeeding and recurrent wheeze at 1 year (Supplementary figure 2) and protective associations between breastfeeding and early wheeze or asthma have been described elsewhere^109,110^. However, we wished to identify any positive or negative associations with human milk components. In a subset of breastfed children with breast milk fatty acid data available (n=966; 110 with asthma, 856 without), increasing percentages of three metabolically linked human milk omega-6 fatty acids in breast milk were associated with 5-year asthma in our initial ExWAS analysis: adrenic acid (AdA; OR: 1.56, 95% CI: [1.25, 1.94]), gamma linolenic acid (GLA, OR: 2.12, 95% CI: [1.41, 3.17]) and dihomo-gamma linolenic acid (DGLA, OR: 2.90, 95% CI: [1.49, 5.64]; Figure 2b). In another analysis that adjusted for additional factors (infant age at time of milk collection while mutually adjusting AdA, GLA, and DGLA components), a positive association was confirmed between upper quintile concentrations of AdA and 5-year asthma (OR: 1.98, 95% CI: [1.12, 3.49]; Supplementary figure 14).

Given that omega-6 fatty acid levels in human milk have been positively-associated with food allergy in a sex-dependent female manner^44^, and AdA/DGLA levels are associated with maternal obesity and other overweight phenotypes^111^, we stratified by sex and added an additional control for maternal BMI. Female infants exposed to breast milk AdA percentages in the highest quintile had increased odds of asthma at 5 years (OR:2.92, 95% CI:[1.02, 8.32]) and 8 years (OR: 4.81, 95% CI: [1.48, 15.66]), whereas no association was observed in males (Figures 4a, 4b). While some transient, early wheeze phenotypes have been attributed to airway size in boys^112^, AdA may be associated with a more persistent form of asthma that first emerges as recurrent wheeze at age 1 (Supplementary figure 2) and continues up to age 8 based on this study. An additional or complementary explanation may involve sex-specific influences in females that are more pronounced by mid-childhood. While high levels of total omega-6 fatty acids in breast milk have been linked with asthma-like symptoms in infants^113^, to our knowledge this is the first report suggesting a sex-specific association between high levels of breast milk AdA and asthma, and requires further study. Using GSEA, breast milk AdA levels in the upper quartile were associated with increased biomarkers of leukocyte migration and the MyD88-dependent TLR pathway (KEGG:04620), and the leading-edge gene profile (TNF, CCL7, CCL4, CXCL8, IL6, CCL3) was consistent with some children having a T2-low pattern reported in neutrophilic asthma (Table 1, Figure 4c). Note that no association with AdA levels and diet were observed. Given AdA’s links with obesity^114^ and cardiometabolic disease^115^, which have inflammatory-associated components, higher AdA in human milk may serve as a marker of maternal metabolic and/or inflammatory physiology. This underlying physiology, including epigenetic and genetic risk shared with the child, could be the real driver of the observed childhood asthma association.

**Fig. 4.**
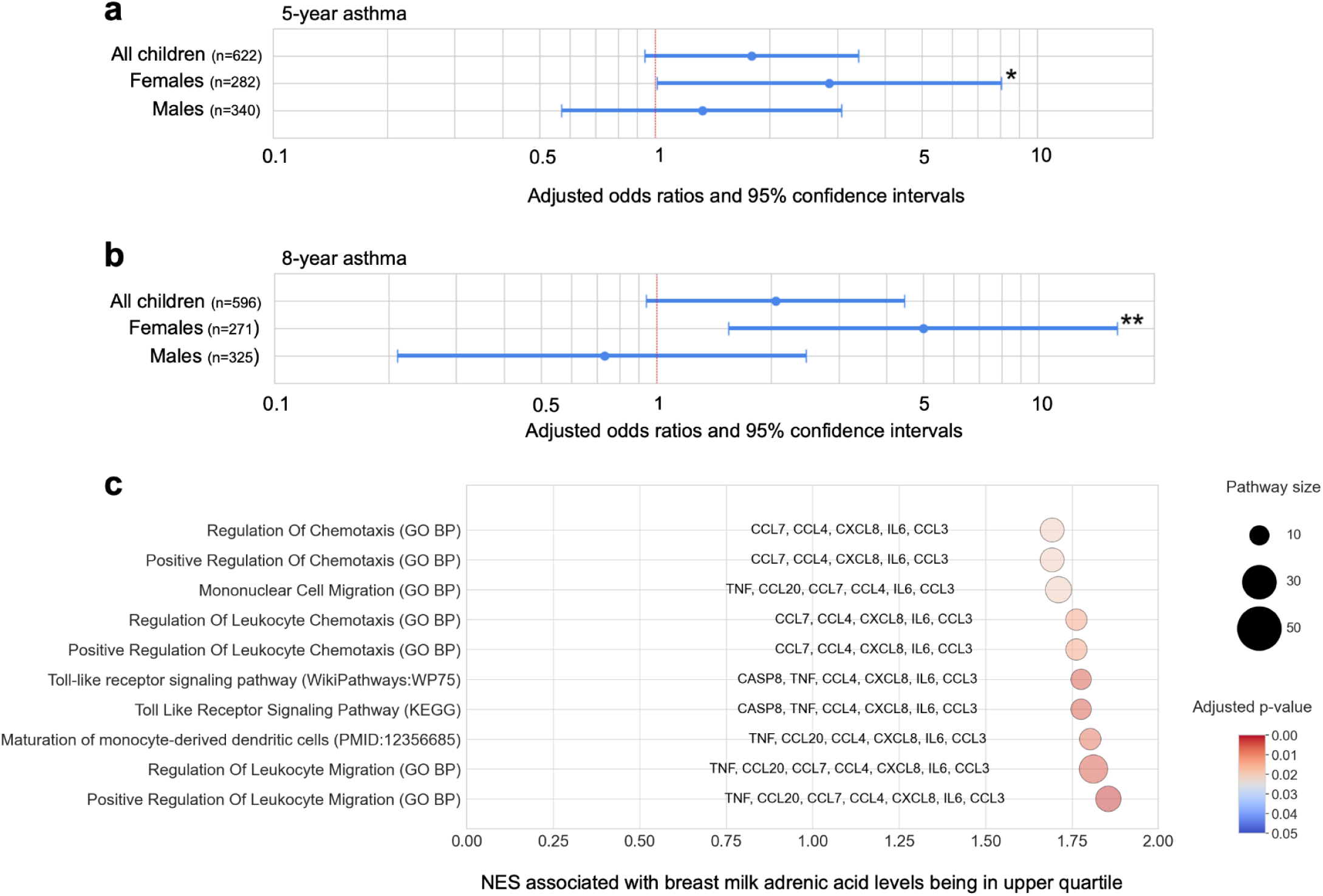
Breast milk AdA levels in the upper quintile are associated with increased odds of girls being diagnosed with asthma, enrichment of serum biomarkers of leukocyte migration and the Toll-like receptor signaling pathway. **a.** Forest plot of sex-stratified association between adrenic acid (AdA) levels being in the upper quintile and odds of having an asthma diagnosis at age 5. **b.** Sex-stratified association between AdA levels being in the upper quintile and odds of having an asthma diagnosis at age 8. Both logistic regression models were adjusted for study site, season of birth, biological sex, birth mode, exclusive breastfeeding status at 3 months, any exposure to tobacco smoke in the home (prenatal), family asthma history, household income, Caucasian ethnicity, number of antibiotic courses (birth - 1year), breast milk GLA and DGLA levels, infant age at time of milk sample collection, and maternal BMI. P-values: * p <0.05, ** p < 0.01. **c.** Gene set enrichment analysis (GSEA) of biological pathways associated with the children’s 5-year serum cytokine dataset in response to AdA levels being in the upper quartile. Figure shows the top significantly-enriched pathways which are primarily related to regulation of leukocyte migration, Toll-like signaling pathway, and chemotaxis. The positive normalized enrichment scores (NES) indicate upregulation of each pathway in children of mothers who had high AdA proportions in breastmilk. The diameter of each circle indicates the set size - a reflection of the number of proteins in the specific molecular signatures database gene set after filtering out genes not included in the serum cytokine dataset. Core enrichment genes that contribute most to the enrichment signal are listed to the side of each bubble.

### Exposure-associated cytokine profiles highlight immunological heterogeneity in childhood asthma

Although participants were not stratified by asthma diagnosis or corticosteroid treatment status, GSEA identified significantly enriched MSigDB gene sets and immune signatures (including T2-low and T2-low/T1-high profiles) that were significantly associated with early-life exposures (Figure 6; Table 1). Prenatal daily hand sanitizer exposure and higher AdA in human milk were both associated with increased pathway biomarkers characteristic of a T2-low cytokine response (Table 1), even though these exposures are not correlated and were independently associated with asthma in a logistic regression model containing both.

In contrast, infants given 2 courses of systemic antibiotic between birth and 12 months had a significant decrease in cytokine biomarkers that enriched for G protein coupled receptor binding, antimicrobial humoral immune response, and chemokine signaling. This response was characterized by a T2-low/T1-high subset profile that included decreased CXCL9, CXCL10 and CXCL11 chemokines, suggestive of a slightly different Th1-driven response^116^ (Table 1, Supplementary figure 15) and a well-characterized Th2 chemokine (CXCL5) previously linked to increased neutrophilia and eosinophilia, and bacterial clearance^117,118^.

Collectively, these chemokine profiles are characteristic of a less treatable endotype of asthma dominated by neutrophils, and can range from T2-low (Th17)-associated to T2-low/T1-high (Th1)-associated responses^78^ and corroborates asthma being a heterogeneous condition with multiple etiologies. In contrast, no associations with Th2-high associated cytokines including IL-4 and IL-5, or signs of increased eosinophils at age 5, were observed in this study. This is characteristic of an allergic asthma endotype, amenable to treatment with corticosteroids, and study participants are likely to have less eosinophilia and lower Th2 cytokines at age 5 years if being treated^119,120^. Environmental exposures previously associated with allergic asthma or atopy include house dust mite, pet dander, mouse droppings/urine/dander, German cockroach, rhinovirus infection, and air pollution including NO2, PM2.5, ozone, and DEHP^121^. We observed a positive association between DEHP and increased eosinophils at age 1 year, and GSEA of chemokines from DEHP-exposed infants identified biomarkers of a T2-low profile that were downregulated at age 5 years, concordant with an atopic response, however T2-high-associated chemokines were also downregulated (Table 1).

Although treated allergic asthma would not be expected to retain a strong T2-high signature in this analysis, each asthma-associated exposure nevertheless remained associated with inflammatory responses, particularly T2-low profiles that are often less treatment-responsive. Collectively, these findings suggest that these exposures may contribute to some treatment-refractory asthma-associated inflammation, with subtly different response profiles.

### Gradient boosting–based machine learning identifies additional early-life asthma risk factors and interactions beyond ExWAS

Several established asthma severity factors, including tobacco smoke and traffic-related air pollution, were not statistically significant in our ExWAS.^122^. We hypothesized that they function in combination with other factors, and so we additionally performed machine learning analyses, which complement ExWAS by also identifying exposure combinations predictive of asthma.

Previously, models including wheezing status and atopy have been used to effectively predict asthma at 5 years of age for clinician use^43^, but we further hypothesized that a model built using early-life environmental exposures and family health history could be used to also identify asthma-associated exposures. In this case we would not want to include intermediate measures of wheeze and atopy that are highly correlated with the asthma outcome and confound the identification of important early exposures. We constructed a 130-feature training dataset of early-life environmental exposures (those associated with asthma via ExWAS or other well known factors described in the literature), plus early child history including birth chart and weight for age between birth and 1 year, and parental health history (features listed in Supplementary file S6).

A holdout dataset used to test the best performing model had a recall of 0.818, Area Under the Receiver Operating Curve (AUROC) of 0.836 and an Area Under the Precision Recall Curve (AUPRC) of 0.161. This is consistent with multiple factors potentially causing asthma, with low precision due to a likely stochastic component to asthma development (for example, high cleaning product exposure would not definitively cause asthma, but rather it would increase the risk of epigenetic changes that impact asthma associated pathways). Determination of the top features contributing to the predictions was assessed by calculating SHAP (SHapley Additive exPlanations) values, which have the added benefit of providing insight into the direction of effect (Figure 5a). Top features included DEHP levels in house dust, family asthma history, father history of atopic dermatitis, use of antibiotics, and use of chemical hand cleaner and hand sanitizer by the mother during pregnancy (all found in the ExWAS), plus additional features linked to primary home residence including pollution levels (several involving PM2.5 but also O3 and NO2), supporting evidence that air pollution can play a role in asthma. To explore this further, SHAP interaction values indicating any combined effect of two features, after accounting for their individual effects, were calculated.The highest ranking ones included interactions with DEHP or a family history of asthma (Figure 5b), with two-dimensional partial dependence plots suggesting that exposure to higher levels of air pollution, particularly PM2.5, but also prenatal exposure to O3 and NO2, can enhance associations between a family history of asthma and odds of the child developing asthma (Figure 5c). There were interactions between family history of asthma and prenatal cleaner/disinfectant use including chemical hand cleaner and hand sanitizer (both assessed at 18 weeks; Figure 5c), and an interaction between higher prenatal levels of O3 and use of antibiotics, that all increased the odds of developing asthma at age 5 (Supplementary figure 16). The latter observation is notable because another study that showed ozone-induced changes in mouse serum metabolomes included metabolites capable of influencing pulmonary hyper-responsiveness, which could be altered in mice that had microbiomes depleted using antibiotics^123^.

**Figure 5.**
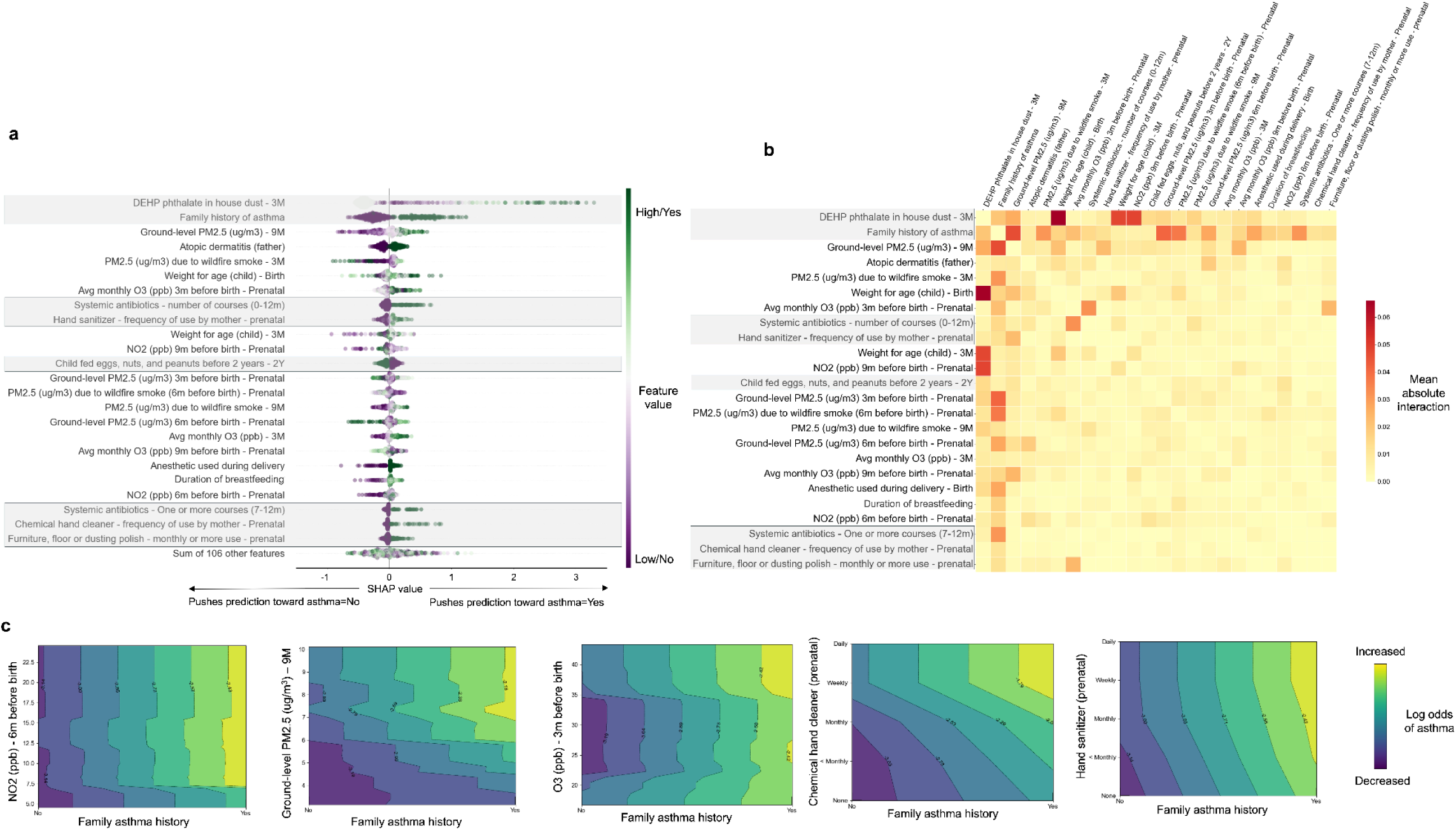
Contribution of individual features, and their interactions for prediction of 5-year asthma using a gradient boosting classifier. **a.** Plot of top features contributing to prediction based on SHAP values (note features are similar to what were returned using a permutation approach, however, SHAP provides insight into direction of effect). Purple dots represent individuals with a lower value (or no response) while green represent individuals with a higher value (or yes response) for that feature. Dots on the right of the baseline indicate that the value pushed the prediction toward asthma=yes, while dots on the left of baseline indicate value pushed prediction toward asthma=no. Grey shaded boxes indicate features that were important in the gradient boosting classifier model and also a significant factor identified by ExWAS. **b**. SHAP interaction heatmap showing mean absolute interaction between features with top SHAP values. **c**. Two-dimensional partial dependence plots of select interactions between family history of asthma, and air quality or prenatal cleaners, that increase the odds of developing asthma at age 5. From left to right: i. Average monthly NO2 (ppb) at 6m prior to birth, ground level PM2.5 (ug/m3) at 9 months post birth, iii. Average monthly O3 (ppb) at 3 months prior to birth, iv. Frequency of chemical hand cleaner use by mother during pregnancy (reported at 18th week), v. Frequency of hand sanitizer use by mother during pregnancy (reported at 18th week). Colormap represents predicted values on the log-odds scale, where higher values (yellow) indicate increased odds of asthma and lower values (purple) indicate decreased odds.

There was a notable overlap in important features (e.g. DEHP, antibiotic use, prenatal cleaning product use, and family asthma history) identified by the machine learning and ExWAS approaches using different wheeze or asthma endpoints. The ExWAS could miss important features associated with asthma because of their correlation with season of birth and study site, or their interactions with each other. Models like gradient boosting complement logistic regression/ExWAS by using a sequential, greedy approach to identifying multiple “weak learners” that can reduce the overall classification error^124^ while the ExWAS allows you to test exposures independently. The overall performance of the different models is consistent with a stochastic process leading to asthma development. Some factors including family asthma or atopy history may represent genetically inherited factors, or transgenerational epigenetic effects, that increase childhood asthma risk, with effects potentially enhanced through interactions with other factors including air quality. This evidence for interactions may explain inconsistency of results between some studies, and evidence of epigenetic changes associated with cleaning exposures is consistent with a stochastic process resulting in progression to asthma.

### Toward identification of early interventions that reduce/eliminate asthma

Integration of CHILD’s diverse, longitudinal datasets into CHILDdb enabled a more wholistic, less biased approach that (1) supported previously-described exposure associations with asthma (e.g. antibiotic use), (2) illustrated how some exposures appear to involve interactions that exacerbate risk (e.g. air pollution), and (3) identified other novel risk factors (Figure 6). Notably, some of the most significant exposures associated with asthma were assessed as early as the 18th week of pregnancy: several disinfectants/cleaning products used frequently prenatally by the mother were each associated with increased asthma risk years later in the child, even if the mother halted such exposure after birth. These cleaning products had significant associations with epigenetic changes in the cord blood at birth, including in inflammatory genes. These data identify potential early intervention targets that merit further study for asthma prevention (such as avoiding certain cleaning product exposures during pregnancy, including occupational use), in addition to actions already being taken to reduce child antibiotic use that were previously found to have measurable benefits at the population level^17^.

**Fig. 6.**
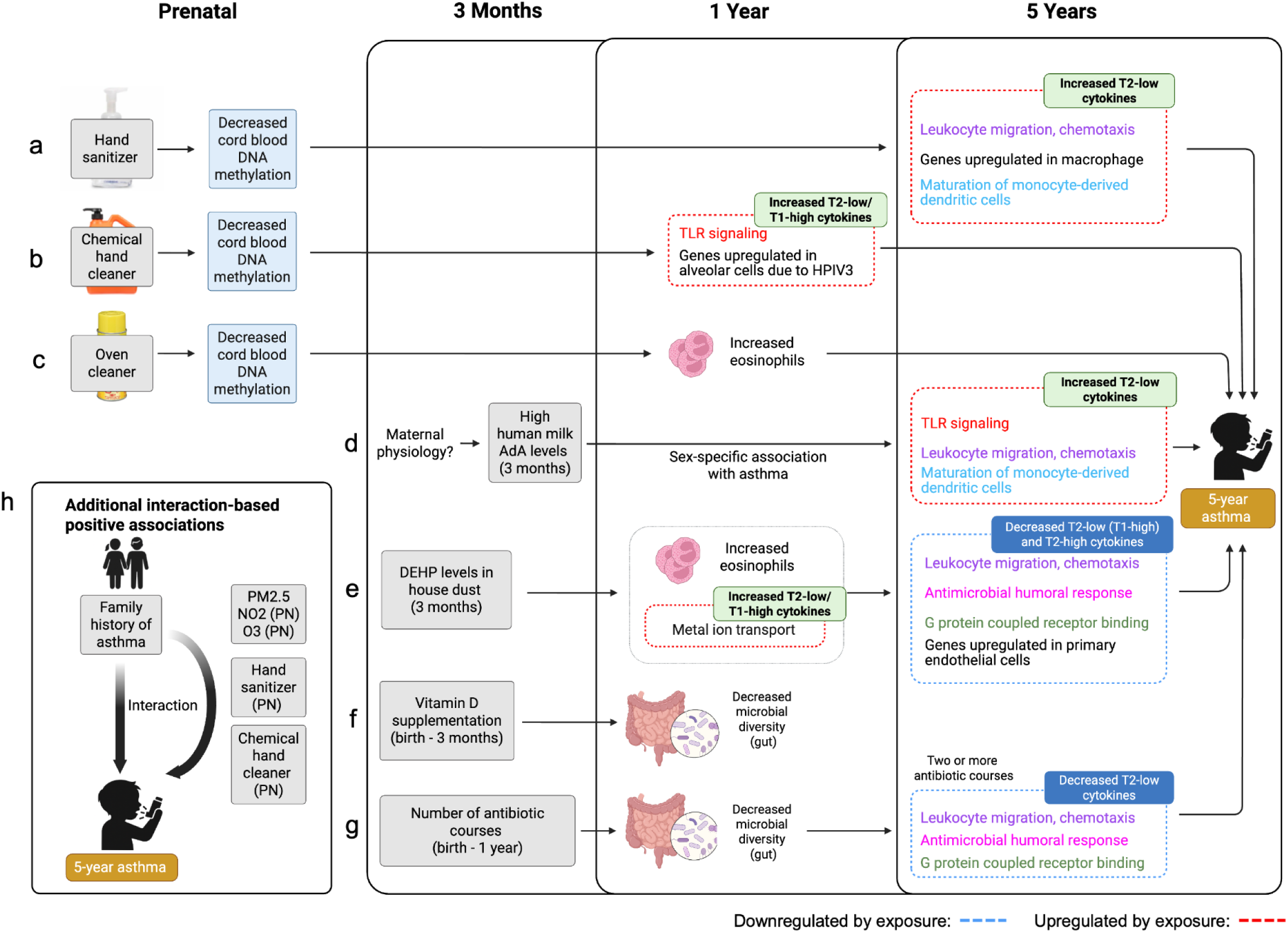
Prenatal and early-life exposures associated with childhood asthma, and associated differences in inflammatory responses and/or microbial dysbiosis, support an etiological heterogeneity for asthma. **a.** Daily use of hand sanitizer, **b.** weekly or more use of chemical hand cleaner, and **c.** weekly or more use of oven cleaner during the prenatal period corresponded with decreased cord blood DNA methylation, which putatively increases transcription of genes contributing to lung inflammation and asthma. Prenatal hand sanitizer was also associated with enrichment of inflammatory pathways characterized by a leading edge subset of T2-low cytokines characteristic of a neutrophilic asthma endotype in a subset of children at 5 years. **d.** Human milk fatty acid proportions may act as biomarkers reflecting maternal physiology. High human milk fatty acid levels, particularly adrenic acid (AdA), were associated with increased odds of asthma, while further analysis demonstrated that AdA proportions in the upper quintile were associated with 5- and 8-year asthma in females. AdA also exhibited increased leukocyte migration, maturation of monocyte-derived dendritic cells, and Toll-Like receptor signalling with a T2-low chemokine profile resembling prenatal maternal daily hand sanitizer use. **e.** Exposure to DEHP concentrations greater than the top quartile in house dust was associated with higher levels of eosinophils and cytokines associated with metal ion transport at age 1. Cytokines associated with leukocyte migration and chemotaxis, G protein coupled receptor binding, antimicrobial humoral response and primary endothelial cell function were downregulated. **f.** Supplementation with vitamin D between birth and 3 months of age was associated with increased odds of developing asthma at 5 years of age, however, the association did not hold up after accounting for additional covariates in a smaller subcohort with complete data for all of the risk factors in this diagram. However, decreased gut microbiome alpha diversity (3 months and 1 year) was associated with vitamin D supplementation and suggests further study is needed. **g.** Early-life antibiotic exposure was linked to asthma and accompanied by decreased microbial diversity in the gut. Children receiving 2 or more courses of antibiotics in the first year of life had decreased levels of T2-low cytokines that may be associated with an emerging T2-low neutrophilic asthma subgroup. Like DEHP, there was a negative enrichment of leukocyte migration/chemotaxis, antimicrobial humoral response and G protein coupled receptor binding pathways. **h.** Additional positive associations with asthma were observed that only appeared via interactions with other factors. For example, family history of asthma or allergy, which can represent an inherited risk factor (genetically or epigenetically), appears to interact with other environmental exposures, including pollution measures like PM2.5, NO2 and O3, to increase the risk of developing asthma. Abbreviations: PN (prenatal), AdA (adrenic acid). Created in BioRender. Winsor, G. (2026) https://BioRender.com/q4sdbbk.

One must be careful though about inferring causation, as for example hospital workers with frequent hand sanitizer use, or house cleaners, may be exposed to other strong cleaners/disinfectants in such an environment that are the true causative agent. Later antibiotic use after 7 months, but not birth to 6 months, was associated with asthma, which could be a mix of causation and reverse causation (those with asthma may have more respiratory infections and require more antibiotics). The phthalate DEHP, which also correlates with other risk factors for asthma including atmospheric NO2 and frequent oven cleaner use, may be both a causative factor, as supported by other mechanistic studies^125,126^, as well as a marker for a lifestyle involving a collection of exposures. Breastfeeding is protective against early childhood wheeze, and high human milk AdA levels have been associated with different metabolic conditions that can be proinflammatory in the mother, so high AdA could also be a marker of genetic/epigenetic, or mother’s inflammatory condition, associated with her child’s more persistent asthma. However, antibiotic use, some cleaning product use, DEHP phthalate exposure, and breast milk components were all significantly associated with asthma in our ExWAS, including after accounting for each other. In addition, they were factors of importance in the complementary machine learning-based analyses.

Linked data from CHILDdb, including epigenetic, inflammatory, and microbiome profiles, enabled us to propose mechanistic hypotheses by which early exposures may drive asthma development, potentially through more than one distinct biological pathway. For example, prenatal hand sanitizer use by the mother may result in epigenetic changes, including in asthma-relevant genes, leading to increased risk of less-treatable neutrophilic asthma at age 5 in some children (Figure 7). The inflammation itself can impact the epigenome, with further potential transgenerational effects, including those demonstrated experimentally in mice, that may in part explain the family history association with asthma^93,127,128^.

**Fig. 7.**
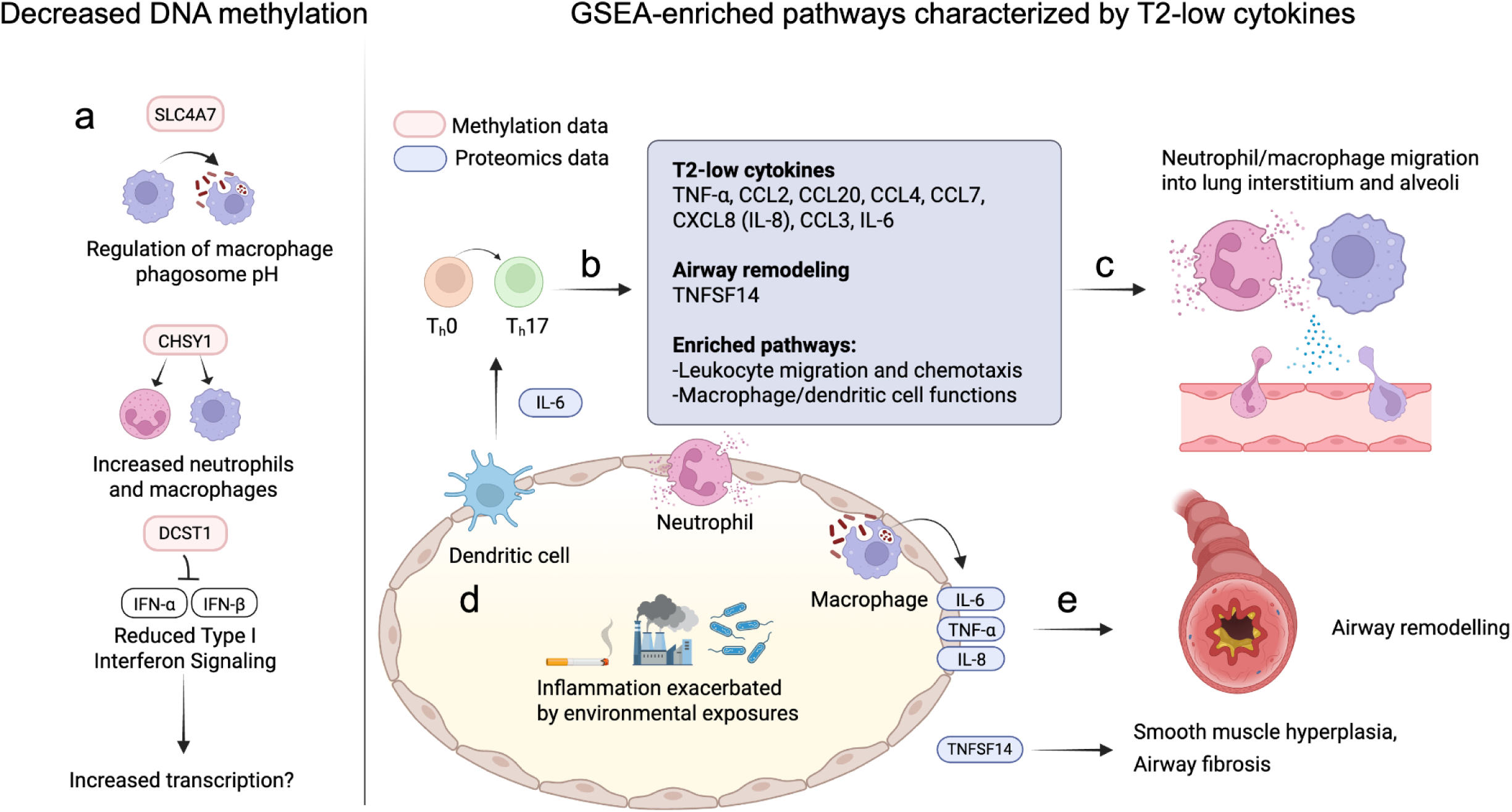
Combining ExWAS, DNA methylation, and Gene Set Enrichment Analysis (GSEA), while drawing on established immunological and asthma concepts, facilitates construction of an example mechanistic hypothesis. In this case an example explaining the association between prenatal hand sanitizer and development of a less-treatable neutrophilic asthma in some children is presented. **a.** Daily prenatal use of hand sanitizer is associated with decreased methylation in cord blood at sites within SLC47 CHSY1, and DCST1 and these genes have been linked to regulation of macrophage phagosome function, increased numbers of neutrophils and macrophages, and associations with negative regulation of Type I Interferon Signaling, respectively. Decreased methylation is putatively accompanied by their increased transcription leading to inflammation and a neutrophilic asthma phenotype in more severe cases. **b.** Increased levels of 5-year serum biomarkers for leukocyte migration and chemotaxis, macrophage, and dendritic cell functions (blue box) have been linked to a T2-low asthma endotype response that is often associated with dendritic cell induction of naive CD4+ T cells into Th17 cells. **c.** Elevated T2-low cytokines lead to increased migration of neutrophils and macrophage migration into the lung interstitium and alveoli. **d.** Migration of neutrophils and macrophages into the alveoli contribute to inflammation which is further exacerbated by common environmental exposures associated with T2-low cytokines. **e.** Uncontrolled inflammation continues to the point where smooth muscle hyperplasia and airway fibrosis (further regulated by cytokines like TNFSF14) lead to airway remodelling in those that have progressed to being diagnosed with asthma. Note that collectively this would be in addition to possible asthma endotypes that would not show up in corticosteroid-treated children. Created in BioRender. Winsor, G. (2026) https://BioRender.com/hlgyy4v.

Notably, both cleaning product use and antibiotic use can impact the microbiome^129,130^, and changes in microbial diversity have been linked to development of childhood asthma^17^, supporting the importance of the microbiome in health and asthmatic disease. In addition, this work further supports the role of air pollution in exacerbating asthma (Figure 5) with smoking more associated with earlier recurrent wheeze conditions at age 1 after controlling for covariates (Supplementary figure 2).

Multiple, independent asthma-associated exposures were associated with subtly distinct inflammatory cytokine pathway responses, including those associated with less treatable asthma (Figure 6; Table 1) - implying that (1) in some children these exposures are associated with a more persistent, less treatable disease burden, and (2) the different exposures result in some heterogeneity in disease response.

There are limitations to our study: The number of serum samples that had cytokine biomarkers assessed limited statistical power for examining how factors like corticosteroid use may impact which inflammatory pathways were enriched according to the GSEA. This work also focuses on one cohort, though the ability of this very richly phenotyped cohort to address so many cofounders and health outcomes is a strength, as well as the cohort’s representation of Canadian population demographic diversity over four harmonized sites. This work also validates other smaller studies in other cohorts, or experimental findings. It has implications for occupational health policy regarding exposures of pregnant women, as well as child exposures including the plasticizer DEHP, that need further study. CHILDdb, through harmonized integration of such diverse data, is a resource that enables these insights and in the future can aid other integrative exposomic investigations of child health and disease.

## Data and code availability

CHILDdb code is freely available under an open source licence. Data described in the manuscript are available by registration to the CHILD database https://childdb.ca and the submission of a formal request. More information about data access for the CHILD Cohort Study can be found at https://childcohort.ca/for-researchers/data-access. Researchers interested in accessing CHILD Cohort Study data for their research should contact.

## Acknowledgements

We thank the CHILD Cohort Study (CHILD) participant families for their dedication and commitment to advancing health research. We thank the whole CHILD Cohort team, which includes interviewers, nurses, computer and laboratory technicians, clerical workers, research scientists, volunteers, managers and receptionists. Key support for this work was provided by the Schroeder Allergy and Immunology Research Institute, Genome Canada, Genome BC, and CIHR, with additional support by Simon Fraser University and the Digital Research Alliance of Canada.

## Methods

### Cohort Description

The CHILD Cohort Study is a prospective longitudinal birth cohort study that recruited 3,624 women with singleton pregnancies from Vancouver, Edmonton, Manitoba (Winnipeg and Morden/Winkler), and Toronto between 2008 and 2012, from which 3,454 delivered healthy, full-term infants, were eligible to commence the study as part of the General cohort^33^. Of the 3,454 infants eligible at birth, 3,133 remained enrolled in the study at the 8-year time point. Participants were phenotyped for conditions including specialist clinician-diagnosed asthma, multiple wheeze and body mass index (BMI) trajectories, food allergies, allergic rhinitis, atopic dermatitis, lung function, and measures of child and mother mental health and behaviour. Questionnaire and environmental datasets, clinical visits, genetic, microbiome and other omics data have been collected for multiple time points ranging from the 18th week of pregnancy to 13 years and beyond.

Ethical approval for the CHILD Cohort Study, including the oversight of the CHILD biological samples and the CHILD database (CHILDdb), was obtained from the local Research Ethics Board of each study site: the University of British Columbia, the University of Alberta, the University of Manitoba, the Hospital for Sick Children, and McMaster University.

The CHILD Cohort Study emphasizes participant and public involvement in all stages of its research. Research questions are informed and generated based on relevance to participants and their families to study those affected by childhood health conditions. Efforts are made to ensure participants’ involvement is meaningful, comfortable, and aligned with their preferences. Study results are shared with participants through various channels, including newsletters and online platforms. Academic and public health publications also communicate findings. We acknowledge and thank all participant advisers and study participants for their valuable contributions.

### Outcomes measured

A definite diagnosis of asthma was ascertained by an in-study specialist during clinic visits taking place at 3, 5 and 8 years of age^33,131^. Recurrent wheeze (assessed for ages 1, 3 and 5 years) was assigned to children having more than 2 episodes of wheeze in the past 12-month period based on parental responses to child health questionnaires that were administered at multiple time points between 3 months and 5 years of age.

### Measurement of human milk components

Human milk fatty acids were measured by gas liquid chromatography as previously described^132^, and human milk oligosaccharides (HMOs) were measured using high-performance liquid chromatography, also as previously described^133^.

### Exposome-Wide Association Study (ExWAS)

To identify exposures and lifestyle factors associated with asthma and/or recurrent wheeze, we conducted a logistic regression-based exposome-wide association study (ExWAS). The ExWAS evaluated the independent contribution of 2,954 exposure variables to asthma or wheeze outcomes (exposures occurring after a given asthma or wheeze endpoint being tested were excluded). These variables spanned common exposure categories encountered from the 18th week of pregnancy through 5 years of age, as well as parental health history prior to birth and maternal mental health assessed from the 18th week of pregnancy through 5 years postpartum (Supplementary Table 1, Supplementary file S3). The majority of variables (2,258/2,954; 76%) captured exposures occurring during the prenatal period through the first year of life.

Binary variables reflecting “daily”, “weekly or more”, or “monthly or more” use of cleaners, disinfectants, air fresheners, and solvents were derived specifically for this study from ordinal home environment questionnaire responses (more details in supplementary methods). The ExWAS analysis was performed in Python using the StatsModels module^134^ where two rounds of logistic regression iterated over the 2,954-variable exposure dataset. The first ExWAS iteration (round 1) tested one independent exposure variable against each of the outcome phenotypes and controlled for study site, season of birth and biological sex of the child. Only participants with complete data for the independent exposure variable and the wheeze/asthma outcome were included (all covariate values were present for all participants in the first round of logistic regression, thus no imputation needed to be performed). The Benjamini-Hochberg method (based on implementation in StatsModels) controlled for false discovery rate (FDR) using a threshold of 0.05. Associations where the FDR < 0.05, the number of asthma or wheeze cases in the exposed group being >=10, and adjusted odds ratio values <0.9 or >1.1 were reported. Significant exposures from the first round of logistic regression were added to a list to be used for a second more stringent round of ExWAS controlling for family history of asthma, number of antibiotic courses between birth and 1 year of age, presence of an older sibling, household income, breastfeeding (exclusive breastfeeding at 6 months of age), birth mode, any exposure to cigarette smoke during pregnancy, study site, season of birth, biological sex and having two Caucasian parents. Only participants with complete data for the independent variable and wheeze/asthma outcome were included in round 2, however missing values for the other covariates were imputed (median for continuous variables and counts, mode for binary, ordinal variables). Columns with missing rows that required imputation included presence of older siblings (8.1% missing), total household income (4% missing), exclusive breastfeeding status at 6 months of age (6.8% missing), birth mode (2.6% missing), prenatal smoke exposure (1.4% missing), and ethnicity (2.2% missing).

Significant associations involving exposures with adjusted odds ratios < 0.9 or > 1.1 and FDR < 0.05, where at least 10 asthma/wheeze cases were exposed, were reported. Stratification by biological sex was also performed for exposures that were significantly associated with asthma at age 5, but were only reported if there was a significant association found in only one of the sexes.

### Ad hoc multivariable logistic regression to validate associations with asthma

A logistic regression using an expanded list of 21 covariates was performed for further validation of significant 5-year asthma associations identified by the original ExWAS analysis. A more stringent model was adjusted for covariates included in the original ExWAS models: family history of asthma, presence of an older sibling, household income, breastfeeding status at 6 months, birth mode, cigarette smoke exposure, study site, season of birth, biological sex, and Caucasian ethnicity (both parents identified as Caucasian). This analysis also adjusted for DEHP (bis(2-ethylhexyl) phthalate) levels in housedust, breast milk fatty acid proportions (AdA, GLA, and DGLA), number of antibiotic courses between birth and 1 year, prenatal maternal exposure to daily hand sanitizer, prenatal maternal weekly or more oven cleaner use, and prenatal maternal weekly or more chemical hand cleaner use, vitamin D supplementation between birth and 3 months, and frequency of oven cleaner use in the home between birth and 3 months. Odds ratios with 95% confidence intervals were estimated and exposures with a p-value < 0.05 were considered statistically significant. Missing values were imputed using the median value.

### Measurement and analysis of cytokine levels

The Olink Target 96 Inflammation Panel (Thermo Fisher Scientific, Waltham, MA) was used to assay 92 protein biomarkers in serum collected from 1- and 5-year-old CHILD participants. Concentrations were analyzed in Olink’s Normalized Protein eXpression (NPX) unit (Thermo Fisher Scientific, Waltham, MA). Exposure variables chosen for Olink analysis were selected from those that were significantly-associated with 5-year asthma in the ExWAS. Continuous variables were converted to a binary variable reflecting the value being in/not in the upper 75th percentile. For the number of systemic antibiotic courses taken between birth and 1 year of age, differences in cytokine levels for serum samples from participants that had taken zero versus one, zero versus one or more, and zero or one versus two courses of antibiotics were assessed. Comparison of levels between participants that had received 3 or more antibiotic courses versus those that did not was not performed because the number of samples from the exposed group was less than 10.

Differences in median between exposure groups were assessed using a non-parametric Mann-Whitney test and corrected for multiple testing using the Benjamini-Hochberg method with FDR<0.05. For select exposures, the OlinkAnalyze version 3.8.2) package^135^ in R was used to perform a gene set enrichment analysis using the olink_pathway_enrichment function while specifying the “GSEA” method, a search against the Molecular Signatures (MSigDB) Database^136^ and providing a dataframe of results from a two-sided t-test performed using the olink_ttest function. The top significant enriched pathways for each exposure, following correction for multiple testing using the Benjamini-Hochberg method with adjusted p-value (more conservative than q value) < 0.05 were reported.

### Characterization of leading-edge cytokines

Each enriched pathway from the GSEA had a list of core enrichment biomarkers (leading edge genes) that contributed most to the enrichment score and are ordered relative to magnitude of change between the two conditions. For each exposure, the list of enriched pathways were collated and core enrichment biomarkers found in at least 50% of the pathways were retained. These biomarkers were manually-curated with citations supporting known associations with asthma, particularly asthma endotypes including T2-high, T2-low, and T1-high – an emerging subset of T2-low asthma (Supplementary table 3).

### Stool sample collection

Sample collection and sequencing were performed as previously described^137,138^. Specifically, stool samples from diapers were collected at a home visit at around 3 months [mean (SD), 3.8 (1.1) months] and a clinic visit at around 1 year [mean (SD), 12.5 (1.6) months]. Samples were aliquoted into four 2-mL cryovials using a stainless steel depyrogenated spatula and were frozen at −80°C.

### Shotgun metagenomic sequencing

Shotgun metagenomic sequencing data were generated by Diversigen (Minneapolis, MN, USA) from fecal samples (average depth of 4.85 million reads per sample, SD[1.79 million]) ^138^ as previously described^138^. DNA was extracted from samples using the MO BIO PowerSoil Pro with bead beating in 0.1mm glass bead plates, with high-quality input DNA verified using Quant-iT PicoGreen. Libraries were prepared and sequenced on an Illumina NextSeq using single-end 1 x 150 reads. Low-quality (Q-Score<30) and length (<50) sequences were removed, and adapter sequences were trimmed. Host reads were removed, and 2,846 samples (1,425 from 3-month visit and 1,421 from 1-year visit) with at least 1 million remaining reads or more were retained for downstream analysis. The accession number for the shotgun metagenomic data reported in this paper is BioProject accession (NCBI): PRJNA838575.

Quality filtered fastq files were processed via Humann3 (version 3.5) and Metaphlan3 (version 3.0.13) to produce functional profiles and taxonomic classifications, respectively, using the Chocophan database (v201901b)^139^. Species represented in fewer than 10% of total samples or less than 0.01% of total abundance were removed, resulting in the retention of 73 species.

### Analysis of gut microbiota

For 3-month and 1-year stool samples, Shannon and Inverse Simpson diversity were calculated using the microbiome R package^140^ while Chao1 diversity was calculated using Phyloseq^141^. Given the degree of variation in collection time for the 3-month and 1-year events, samples collected between 2.5 and 4 months of age were retained for the 3-month analysis while samples collected between 11 and 13 months of age were retained for the 1-year analysis. In order to test for associations between asthma- and wheeze-associated exposures and alpha diversity, mixed linear modelling was conducted using the statsmodels (version 0.12.1) mixed_linear_model module^134^. For each regression performed, study site was added as a random effect, while breastfeeding status, delivery mode, presence of an older sibling, family asthma history, use of systemic antibiotics, sample processing time (time between collection and deep freezing the aliquots), and exact age of stool sample collection (weeks) were added as covariates. False discovery rate was controlled by adjusting P-values using the Benjamin-Hochberg method.

Relative species abundances were log-transformed, and the “Maaslin2” package was used to perform linear mixed-effects modelled (MaAsLin2 function)^142^ to identify associations with prenatal cleaning product and hand sanitizer use while specifying study center location as a random effect and adjusting for stool sample of collection age, genetic ancestry of infants PC1-PC10^143^, family income at prenatal enrollment, and biological sex. Centered log-ratio transformation (clr) was performed by first adding a pseudocount of one half of the minimum relative abundance value following a clr transformation using the compositions package^144^. To identify associations between species and the “Healthy/Non-Allergic” phenotype, Maaslin2 was used to build linear mixed-effects models with study center location as random effect and adjusting for stool sample collection age (defined as time between collection and deep freezing the aliquots). For vitamin D supplementation, the Maaslin3 package^145^ was used to identify associations with both abundance and prevalence of species with study center location as random effect while adjusting for birth mode, exclusive breastfeeding, older children in home, systemic antibiotic use, age of infant, and sample processing time. Only features that had an FDR <0.05 and an absolute coefficient value greater than 0.2 were retained for plotting in the heatmap. The heatmap was created using the complexHeatmap package^146^.

### Microarray genome-wide DNA methylation profiling

DNA was extracted from cord blood samples using the DNeasy Blood & Tissue Kit (Qiagen, Venlo, The Netherlands), samples were bisulfite converted using EZ-96 DNA Methylation kit (Zymo Research, Irvine, CA,) and DNA methylation profiles of the samples were measured with the Infinium MethylationEPC BeadChip array (EPIC) (Illumina, San Diego, CA). Raw intensity IDAT files for 866,836 data points encompassing 863,904 CpG sites were produced and subsequently preprocessed for downstream analysis.

### DNA methylation preprocessing and data reduction

All available cord blood DNA methylation data (n = 838) were processed in RStudio (version 4.0.3) as previously described^147,148^. Briefly, the ewastools package^149^ was employed for sample quality control to assess the technical parameters, the minfi package was used to assess intensities and check sex concordance and the lumi package was used to detect potential outliers^150–152^. Probe filtering was performed based on the Pidsley annotation to remove cross hybridizing and polymorphic probes, and BMIQ normalization was done in conjunction with noob to account for probe type bias and background correction^153,154^. Fifteen of the 838 samples were removed due to failure of one or more quality metrics. Finally, to better account for known technical confounding factors, variation associated with batch (i.e., chip and row) was assessed and removed using the ComBat function from the sva package^155^. After all preprocessing steps, a total of 813 samples and 786,363 probes were included in the cleaned sample set. For further data reduction, DNA methylation probes were subjected to interquartile range filtering to subset only variable probes where the DNA methylation β value varied by at least 5% across samples in the 95th percentile^156,157^. A total of 356,037 variable probes were included in the Epigenome-wide association analysis. Of the 813 samples included in the DNA methylation data set, a further subset of 808 children had data regarding exposure to the cleaning variables of interest. A robust linear regression model using Huber M-estimation in the MASS package was used for site-by-site epigenome-wide association analysis^158^. The following formula was employed for each exposure variable:

~~~
CpG ∼ [Exposure Variable] + Biological sex + Gestational age + Birth mode
+ Genetic ancestry principal components + Cell type principal components + ε
~~~

Biological characteristic covariates known to contribute significantly to DNA methylation differences, i.e., sex, gestational age, genetic ancestry, birth mode, and cell type proportions were included in the models^159^. CpG sites were determined to be significant if they met a Benjamini–Hochberg adjusted p-value threshold of < 0.05 and had a delta beta greater than 3%.

### Prediction of childhood asthma and identification of most important features using machine learning analyses

The GradientBoostingClassifier algorithm from Scikit-learn^160^ in Python (version 1.8) was used to predict 5- year asthma cases using a training dataset of 130 features including early environmental exposures, parental health history (including asthma, atopy and mental health), child birth, chart and child anthropometric data (weight for age at birth, 3 months, and 1 year). In order to avoid using variables that reflect early manifestations of asthma that could potentially dominate the models, we purposely excluded variables describing children’s wheeze, asthma and allergy.

We included a baseline variable dataset used previously to predict definite cases of 5-year asthma based on specialist clinician-diagnosis ^43^. This baseline dataset also included significant variables from the round 1 ExWAS analysis where the FDR cutoff was relaxed to 0.1, and a dataset of other variables linked previously in scientific literature to childhood asthma, including CANUE variables linked to the postal codes of primary residences (Supplementary file S6).

Prior to building the model, all features were confirmed to have Spearman rank correlations < 0.9 with each other. Participants containing contradictory parent asthma histories based on responses in 2 separate questionnaires were dropped. Participants where either the 3-year or 5-year asthma diagnosis were missing were also dropped, as well as any participants where 3-year or 5-year asthma was classified as ‘probable’ but not ‘definite’ based on criteria defined previously^33^. The mother’s previous number of pregnancies, duration of stay in hospital after the child was born, and duration of breastfeeding were transformed using the natural logarithm of one plus the input array in order to compress their distributions. Discrete variables were created from continuous data representing mother mental health Perceived Stress Scale^161^ and Center for Epidemiologic Studies Depression Scale^162^). No further processing was performed on the other features prior to generating the training and validations sets.

The outcome variable was 5-year asthma based on diagnosis by a specialist clinician and the dataset contained 1672 participants without asthma and 108 participants for the 5-year endpoint. Data was split into 70% training / 30% validation (holdout) datasets. The training set underwent stratified 3-fold cross-validation with missing values in each fold being imputed using mode for categorical variables and median for continuous variables. The gradient boosting classifier used negative log loss for internal optimization and hyperparameters included class_weight = balanced, learning_rate=0.05, max_depth=7, max_iter=100, and min_samples_leaf=25. Cross validation of models was assessed using negative log loss and an optimal decision threshold based on a target recall of 0.8 applied in order to prioritize identification of true positive asthma predictions. Recall, AUROC and AUPRC were calculated for the holdout dataset and SHAP (SHapley Additive exPlanations) values^163^ were calculated for the holdout dataset using the shap package (version 0.46.0) in Python. SHAP interaction values^164^ were calculated for the top 20 features using the shap package and the resulting mean absolute interaction for each pair was plotted in a heatmap.

