## Supplementary material for "Exposomics for childhood asthma"

### Supplementary results

#### Low between-category correlation of asthma-associated exposures supports a multifactorial etiology for asthma

The diverse CHILDdb data allows us to further examine the correlation network of associations involving exposures found to be significant. This, coupled with a model including all exposures together (plus known confounders), can support identification of more probable causal factors.

An all-versus-all correlation analysis, of significant 5- and 8-year doctor diagnosed asthma-associated factors identified by ExWAS, with covariates, was visualized using a correlation globe[165] (minus those likely due to reverse causation; Supplementary figure 4a, Supplementary file S5). There was a distinct lack of correlations between factors belonging to the same exposure "category", as labelled in Supplementary figure 4a (e.g., the breast milk fatty acids as an exposure category all correlated with each other, but did not correlate with other categories, such as antibiotic or cleaning product use). The only exceptions were very weak correlations between DEHP phthalate levels in house dust and either the child being given any vitamin or supplement at age 0-3m ($\rho$=0.111), or the mother's frequency of oven cleaner use postnatally 0-3m ($\rho$=0.123). The latter is notable, given oven cleaning spray has previously been positively associated with DEHP and its metabolites in women's urine[166]. Very frequent oven cleaner use is negatively associated with income, and positively associated with other cleaning products like tile cleaner and chemical hand cleaner – consistent with house cleaning and related occupations (Supplementary table 4).

Collectively, the general lack of correlation between these different categories of asthma-associated exposures, including after controlling for family asthma history, is consistent with potentially multiple causative factors and exposure pathways that increase risk of asthma - even if they may all converge on the same or similar eventual biological/inflammatory process. However, to better test this, a logistic regression model was built including all asthma-associated exposures together (plus known confounders), of a subcohort of participants without imputation. Human milk fatty acid adrenic acid continued to be significantly associated with asthma, as did antibiotic use, DEHP Phthalate, and multiple prenatal cleaning product exposures (Supplementary figure 4b). Due to the smaller sample size in this subcohort (n=549), it cannot be ruled out that other exposures identified in the initial ExWAS are not still significant, however, the observation of multiple different types of asthma-associated exposures, that themselves do not correlate well with each other, and are still significant after controlling for each other, is consistent with a multi-causal etiology for asthma.

In contrast, the initial association between infants being given vitamins or supplements between birth to 3 months, and asthma, was no longer statistically significant after correction for DEHP levels (which it weakly correlates with; Figure 4; Note vitamin use at this age is 99% vitamin D; Supplementary Figure 13). However, this vitamin association with asthma needs more study, since, unlike DEHP, Vitamin D supplementation was associated with decreased diversity of the gut microbiome, including significantly lower 3-month alpha diversity scores (Supplementary figure 13a), including when stratified based on exclusive breastfeeding status, with linear mixed effects modeling adjusting for multiple covariates known to influence both asthma and gut microbiota (Supplementary figure 13b). A separate CHILD study using a different 16S gut microbiome dataset observed microbiome changes if mothers took prenatal and postnatal vitamin D supplements[84]. Collectively, these analyses suggest that while the impact of infant Vitamin D supplementation on their gut microbiome needs more study, the associated phthalate DEHP exposure is of primary note, given its association with asthma even after correcting for so many confounders.

The different primary categories of asthma-associated factors identified (Figure 3a) were selected for more in-depth investigation as described below, including cytokine, microbiome and/or epigenetic changes associated with these exposures (shaded exposures in Figure 3a). Notably, DEHP, antibiotic, and cleaning products investigated were associated with serum cytokine changes and other inflammatory markers (such as higher eosinophil/lymphocyte ratios), consistent with their association with asthma (Supplementary figures 6, 7, 8, 9, and 10 and described further below).

#### Prenatal cleaning product and hand sanitizer use is significantly associated with asthma following correction for use of other cleaning products and later time points

To address potential postnatal confounders for the prenatal exposure, logistic regression controlling for any postnatal use of the same product was performed for the 5-year asthma analysis, and still supported that prenatal use of all three cleaning/disinfectant products examined remained positively associated with asthma (Supplementary figure 5a). The possibility that use of other correlated cleaning products during pregnancy could be confounding their association with asthma was also considered. Using weekly prenatal oven cleaner, chemical hand cleaner, tile cleaner, and daily hand sanitizer as additional covariates for the initial ExWAS, logistic regression confirmed the positive association between each individual cleaner and asthma at 5 years (Supplementary figure 5b).

### Supplementary methods

#### Maternal/infant vitamin D use and logistic regression

Maternal vitamin D supplementation during pregnancy was assessed by a questionnaire administered at the 18-week prenatal period asking “How often did you take vitamin D?” with options Never, < 1 per month, 1-3 times per month, 1-3 times per week, 4-6 times per week, and every day. For mothers who took vitamin D 1-3 times per week or more, a binary variable indicating maternal vitamin D use was derived. Maternal multivitamin use during pregnancy was assessed by the same questionnaire where mothers were asked “How often did you take multivitamins?” with options Never, < 1 per month, 1-3 times per month, 1-3 times per week, 4-6 times per week, and every day. For mothers who took multivitamins 1-3 times per week or more, a binary variable indicating maternal multivitamin use was derived. Infant supplementation with vitamin D between birth and 3 months was assessed on a nutrition questionnaire administered at the 3-month visit that asked if the infant was given vitamin D as D-Vi-Sol D-Drops, Poly-Vi-Sol Iron, Tri-Vi-Sol A-C-D, Poly-Vi-Sol A-C-D-B1-B2-B3-B6, as a source mentioned under “other” on the nutrition questionnaire, or explicitly mentioned on the medication questionnaire. Maternal multivitamin use during pregnancy was assessed by the same questionnaire where mothers were asked “How often did you take multivitamins?” with options Never, < 1 per month, 1-3 times per month, 1-3 times per week, 4-6 times per week, and every day. For mothers who took multivitamins 1-3 times per week or more, a binary variable indicating maternal multivitamin use was derived.

Logistic regression was performed to look for association between infant vitamin D supplementation and 5-year asthma where participants were stratified by breastfeeding status (exclusive, partial, zero) with exclusive breastfeeding as the reference, or exclusive breastfeeding status (yes, no), where participants with missing data were removed. Another logistic regression stratified participants by maternal vitamin D supplementation with or without a multivitamin (yes/no) during pregnancy and the vitamin D association with 5-year asthma was modelled using 12 covariates (listed in supplementary figure). Participants with any missing data were dropped. If the mother took vitamin D prenatally, and the infant was given vitamin D between birth and 3 months, a variable indicating this pattern of use was derived and used for an all-in logistic regression analysis looking for associations between environmental exposures used by the ExWAS (described above) where exposures taking place prenatally and up to 3 months of age (1631 exposures), and homes where this pattern of vitamin D use was a practice. For each logistic regression performed, adjustments were made for study site, season of birth, caucasian ethnicity, exclusive breastfeeding status, family income and mother's education. P-values were adjusted for false discovery rate by using the Benjamini-Hochberg method. Significant associations and odds ratios that were >= 3.0 or <=0.7 were shown in a figure, while a file containing all significant exposures was stored in an Excel file.

#### Analysis of proportions of human milk components

Breast milk samples were collected at approximately 3 months of age, therefore children breastfed less than 3 months were excluded from the analysis. For the ExWAS analysis, log-transformed relative breast milk fatty acid, human milk oligosaccharide, and endocrine hormone levels (proportion of total) were used. For subsequent logistic regression analysis of associations with 5-year asthma, breast milk fatty acids were dichotomized at the 75th percentile to distinguish infants with notably elevated exposure without assuming a linear relationship across the full range of levels. . A logistic regression containing the fatty acids GLA, DGLA, and AdA was performed to test for association with 5-year asthma while adjusting for age of infant at time of milk collection, Caucasian ethnicity, family asthma history, biological sex, number of systemic antibiotic courses (birth-1y), prenatal smoke exposure, study site, household income, and birth mode. Participants were then stratified by sex, and an additional logistic regression was performed to test the AdA association with 5-year asthma, while adjusting for the same confounders as above plus BMI of the mothers. Participants with missing data were dropped for the latter two logistic regressions. Odds ratios with 95% confidence intervals were estimated and exposures with a p-value < 0.05 were considered statistically significant.

#### Analysis of phthalate levels

Concentration of phthalates levels in 3-month, 1-year, and 3-urine urine phthalates (MBP, MBzP, MCPP, MEHHP, HEMP, MEOHP, and MEP) and dust phthalates (DEP, DiBP, DNBP, BzBP, and DEHP) are based on sample preparation described previously[167,168]. Measures were log-transformed prior to ExWAS analysis and other downstream analyses. A more stringent logistic regression analysis using StatsModels (version 0.14)[134] tested the association between log-transformed DEHP and 5-year asthma in a subcohort of 549 participants with complete data for DEHP, additional covariates, and 5-year asthma. Additional covariates included season of birth, study site, household income, years of education (mother), prenatal tobacco smoke exposure, older siblings, number of antibiotic courses, biological sex, Caucasian ethnicity, family asthma history, exclusive breastfeeding status, prenatal daily hand sanitizer use, prenatal weekly oven cleaner use, prenatal weekly chemical hand cleaner use, frequency of oven cleaner use (birth-3m), vitamin D use by infant (birth-3m).

#### Analysis of prenatal cleaning product and disinfectant use

On the prenatal 18-week, 3-month, 1-year, 3-year, and 5-year home environment questionnaires, participant families were asked if and how often they used specific cleaning and disinfectant products. Options were ordered on a likert scale and listed as ‘No’, ‘< Monthly’, ‘Monthly’, ‘Weekly’, and ‘Daily’ (For chemical hand cleaner, the question was not asked on 3-year and 5-year questionnaires). Binary variables were derived from these responses in order to represent Monthly or more (yes to monthly, weekly or daily; 0 or 1), Weekly or more (yes to weekly or daily; 0 or 1), or Daily use (0 or 1) of the product. For the prenatal questionnaire only, the questionnaires asked if the mother or somebody else in the home used the product. In this case, derived binary variables reflecting monthly, weekly or daily use by the mother, or by anyone in the household, were generated separately. Since other studies have assessed combined use of multiple cleaning products[91], we derived binary variables indicating use of multiple disinfectant products during pregnancy and up to the first year of life (e.g. “monthly or more use of hand sanitizer, disinfectant, and disinfectant in the bedroom”), and used them in the ExWAS and analysis of DNA methylation profiles.

To identify mothers who reduced their use of daily hand sanitizer during pregnancy to less than daily between birth and 3 months (i.e. weekly or less), an additional binary variable was generated and used for the ExWAS and analysis of DNA methylation profiles.

To assess if prenatal use of other cleaners/disinfectants was confounding the observed associations with 5-year asthma, a single logistic regression was performed. It adjusted for use of prenatal daily hand sanitizer, weekly oven cleaner, weekly chemical hand cleaner,, covariates used in the ExWAS analysis, and weekly prenatal use of tile cleaner (identified as being correlated with the other cleaners).

To rule out any chance that postnatal cleaner use was confounding the prenatal associations with asthma, an additional logistic regression was performed. It tested the association between use of daily hand sanitizer, weekly oven cleaner, and weekly chemical hand cleaner (degreasing agent) by the mother during pregnancy and 5-year asthma while adjusting for any postnatal use of the same cleaning or disinfectant product type (plus additional ExWAS covariates). The postnatal cleaning variables reflected frequency of use of the products reported in the 3-month, 1-year, 3-year and 5-year questionnaires (corresponding to prenatal weekly oven cleaner, prenatal daily hand sanitizer) or 3-month and 1-year questionnaires (corresponding to prenatal weekly chemical hand cleaner, frequency of prenatal chemical hand cleaner).

#### Construction of an exposome correlation globe

Exposures that were significantly-associated with either 5- or 8-year asthma, ExWAS covariates (family history of asthma, presence of an older sibling, household income, exclusive breastfeeding at 6 months of age, any breastfeeding at 3 months of age, birth mode, biological sex and ethnicity), and previously described risk factors referenced in the literature (smoking[61], NO2[55], PM25[19], antibiotic use by mother[169], acetaminophen use by mother[170], and maternal distress[171]) underwent an all-versus-all Spearman rank correlation analysis (scipy.stats, version 1.5.2). For each correlation, at least 100 participants were required after dropping participants with missing values. False discovery rate was controlled for using the Benjamini-Hochberg method with FDR cutoff of $q<0.05$. The remaining pairs of correlations with an absolute value of >= 0.1 were visualized in an exposome correlation globe generated using Circos[165,172]. To provide more insight into other exposures that may be co-occurring with significant exposures identified by ExWAS, and justify the addition of additional covariates to more stringent downstream logistic regression analyses, we performed ad hoc correlation analyses between the following exposures found significant in the initial ExWAS: infant vitamin D supplementation (birth-3m), dust DEHP levels, breast milk fatty acids (GLA, DGLA, AdA), or prenatal cleaner use (hand sanitizer, oven cleaner, chemical hand cleaner), and the other 2983 variables in the starting ExWAS dataset. These correlation tests used the same cutoffs as the correlations performed for the correlation globe analysis.

#### Analysis of white blood cell ratios

Complete blood counts (CBCs) were performed on children's venous, heparanized blood collected at 1- and 5-year time points in outpatient clinics and stored at -80℃ as previously described[137]. Ratios of absolute values for eosinophils, neutrophils, monocytes and lymphocytes were calculated to yield eosinophil/lymphocyte (ELR), neutrophil/lymphocyte ratio (NLR), eosinophil/neutrophil ratio (ENR), and monocyte/lymphocyte ratio (MLR) at 1 and 5 years of age. All ratios underwent natural log transformation and were plotted to assess normality before analysis. Mann-Whitney U tests for differences in median between binary exposure groups were performed and p-values were adjusted for false discovery rate using the Benjamini-Hochberg method. Any statistical differences were further verified using multiple linear regression analysis using statsmodels.formula.ols (Version 0.12.1) to obtain adjusted beta coefficients for the binary exposures tested while adjusting for study site, birth mode, biological sex, caucasian ethnicity, any breastfeeding at 3 months, positive skin prick tests for any food allergens or aeroallergens at 1 year, atopic dermatitis status at 1 year, and family history of asthma. For the multiple linear regression testing, only participants with complete data for the tested exposure, covariates, and outcome (white blood cell ratios) were included.

#### Analysis of mother's occupations during prenatal period

The National Occupational Classification system (NOC) 2021 Version 1.0 was used to convert free-text occupation descriptions provided by the mothers to standard occupation categories. The level three tier was chosen to classify occupations because it provided a good balance between granularity and sufficient statistical power for analysis. Subjects that did not provide a free-text occupation description or where a reliable classification could not be reached were excluded from the analysis. Afterwards, occupation classes falling under the Health Occupation category (except for managers in Healthcare) were grouped together given that these occupations offer health care directly to patients or work in close proximity to them as opposed to managerial occupations and occupations in other sectors. These individuals could have had occupational exposure to hand sanitizer or other cleaning agents.

For comparing the use of different types of cleaners among different groups we performed a chi-square test of proportions followed by a post hoc test using the Benjamini–Hochberg procedure. The results with the adjusted p-value < 0.05 were retained for plotting. The correlation analysis was performed between 2,702 exposure variables and occupation groups, the Spearman Rank correlation was used and multiple testing correction was performed using the Benjamini-Hochberg procedure. Only results that had an FDR < 0.05 an absolute coefficient value greater than 0.1 were retained for plotting in the heatmap.

### Supplementary tables and figures

#### Supplementary figure 1

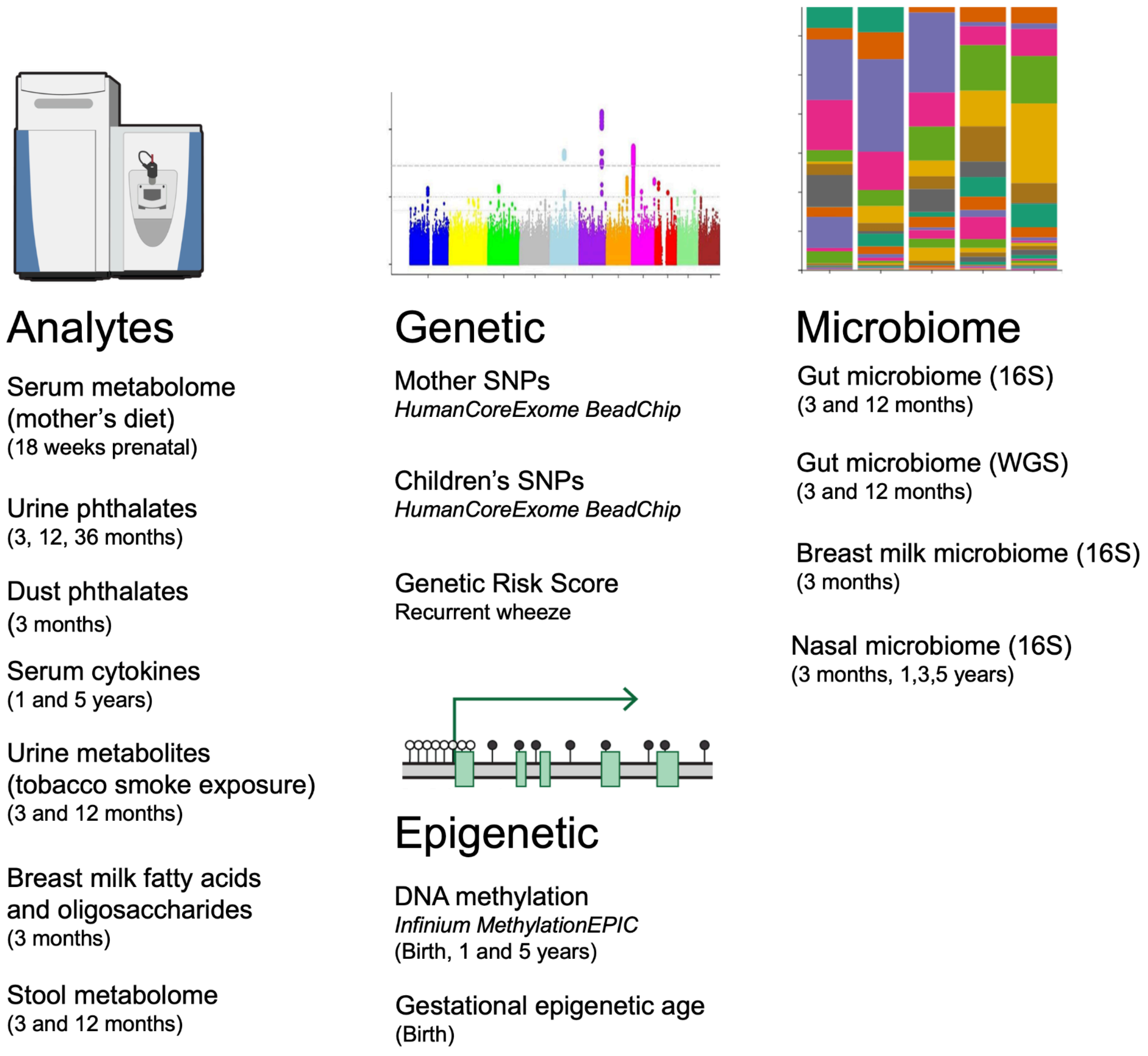


**Supplementary figure 1 | Biological sample-derived data that has been integrated into CHILDdb.**

### Supplementary table 1

**Supplementary table 1 |** Overview of categories and number of variables collected from the 18th week of pregnancy to 8 years of age that were used for the Exposome-Wide Association Study. See also Supplementary file S3 for the full list.

| | **Prenatal** | **Birth** | **3M** | **6M** | **1Y** | **18M** | **2Y** | **30M** | **3Y** | **4Y** | **5Y** | **8Y** |
|---|---|---|---|---|---|---|---|---|---|---|---|---|
| Air quality (CANUE) | 84 | | 23 | 26 | 62 | | | | 8 | | 17 | |
| Birth chart | | 5 | | | | | | | | | | |
| Breastfeeding | | | 2 | 4 | 1 | | | | | | | |
| Built environment (CANUE) | 67 | | 67 | 45 | 41 | | | | 41 | | 73 | |
| Chemical analytes (urine) | | | 14 | | 10 | | | | 8 | | | |
| Chemical analytes measured in breast milk | | | 51 | | | | | | | | | |
| Child Food Frequency | | | | | | | | | 112 | | 114 | |
| Child Health | | | 1 | 1 | 7 | | | | 8 | | 9 | 2 |
| Child Nutrition | | | 65 | 77 | 57 | 35 | 56 | 56 | | | | |
| Climate (CANUE) | 49 | | 50 | | 33 | | | | 10 | | 59 | |
| Father's health | 1 | | | | | | | | | | | |
| Food Packaging and Preparation | | | | | | | | | 17 | 17 | 19 | |
| Greenness (CANUE) | 6 | | 6 | 12 | | | | | | | 6 | |
| Home Environment | 412 | | 444 | 1 | 26 | | | | 10 | | 7 | |
| Medications taken (child) | | | 21 | 20 | 25 | | | | | | | |
| Medications taken (mother) | 3 | | 8 | | | | | | | | | |
| Mother Vitamins and Supplements | 231 | | 52 | | | | | | | | | |
| Mother serum metabolome | 44 | | | | | | | | | | | |
| Mother's health | 14 | | | | | | | | | | | |
| Mother's mental health | 2 | | | 2 | 2 | | | | 2 | | | |
| Neighbourhood-level sociodemographics (CANUE) | 15 | | 15 | 9 | 11 | | | | 11 | | 15 | |
| Parental health | 1 | | | | | | | | | | | |
| Smoking | 1 | | 1 | 1 | 2 | | | | 1 | | 1 | |
| Socioeconomic status | 3 | | | | | | | | | | | |
| UV light (CANUE) | 10 | | 7 | | 8 | | | | | | | |

#### Supplementary table 2

**Supplementary table 2 | Asthma and wheeze outcomes examined in this study.** Number of cases (with percentage of the whole cohort in brackets) of clinician-diagnosed asthma (definite), clinician-diagnosed asthma (definite and possible) and recurrent wheeze (defined as more than 2 wheeze episodes in the past 12 months) - presenting at 1-, 3-,5- and 8-year time points in the CHILD Cohort Study.

| | Number of cases | | | |
|---|---|---|---|---|
| **Outcome** | **1 year** | **3 years** | **5 years** | **8 years** |
| Asthma (definite and possible cases, clinician) | | 361 (13.2%) | 412 (15.6%) | 232 (9.7%) |
| Asthma (definite cases, clinician) | | 167 (6.6%) | 165 (6.9%) | 125 (5.5%) |
| Recurrent wheeze | 249 (7.9%) | 252 (8.8%) | 202 (7.4%) | |

### Supplementary figure 2

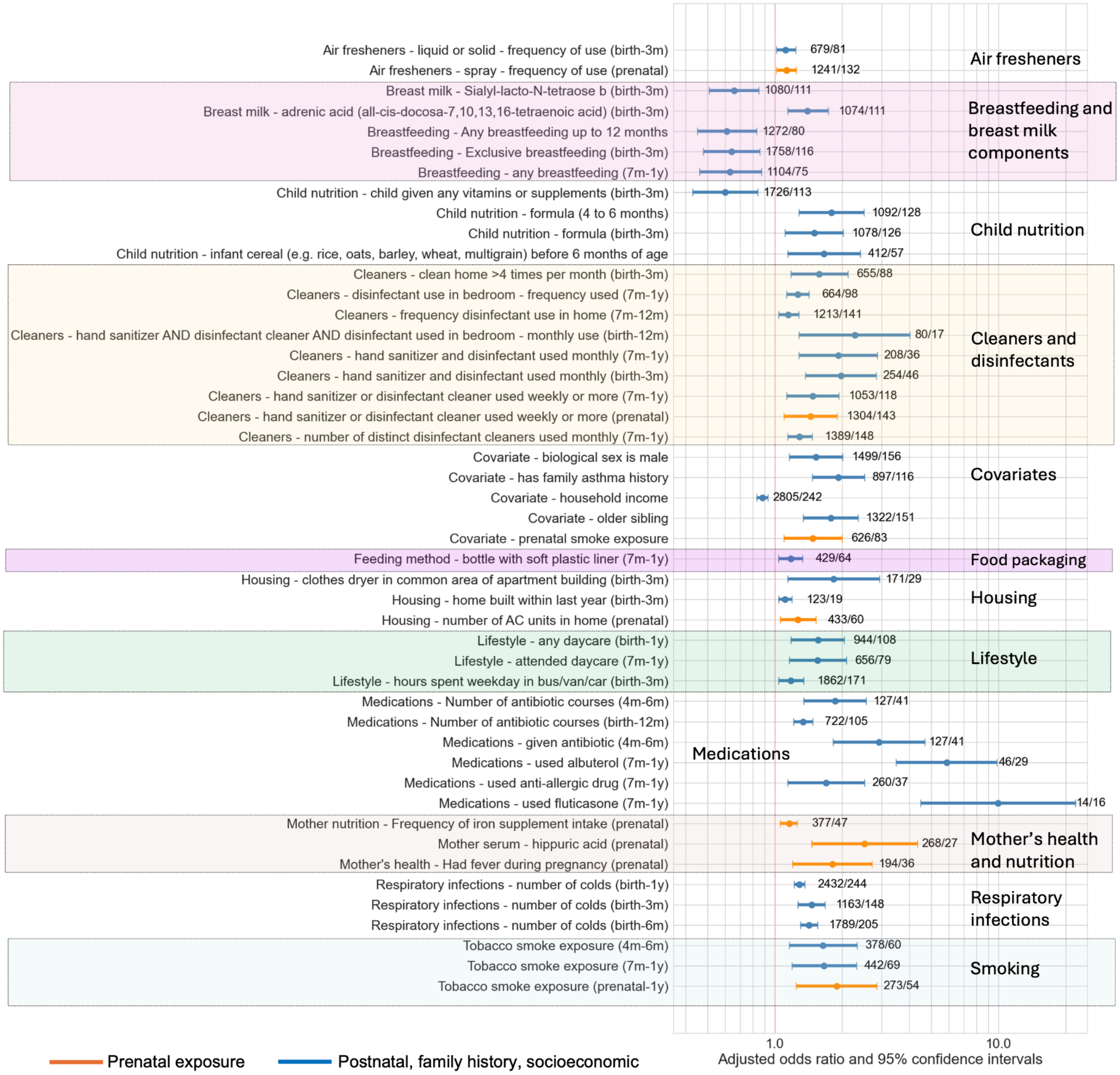

**Supplementary figure 2 | Exposures identified by ExWAS round 2 as being significantly associated with recurrent wheeze at 1 year of age.** 2110 exposures taking place prior to 1 year of age were assessed for associations with 1-year recurrent wheeze. Orange and blue horizontal plots are adjusted odds ratios and 95% confidence intervals for prenatal and postnatal exposures, respectively. Numbers to the right of each plot represent total number non-cases and cases subjected to each exposure, respectively. Major groups of exposures are labelled and coloured for additional clarity.

### Supplementary figure 3

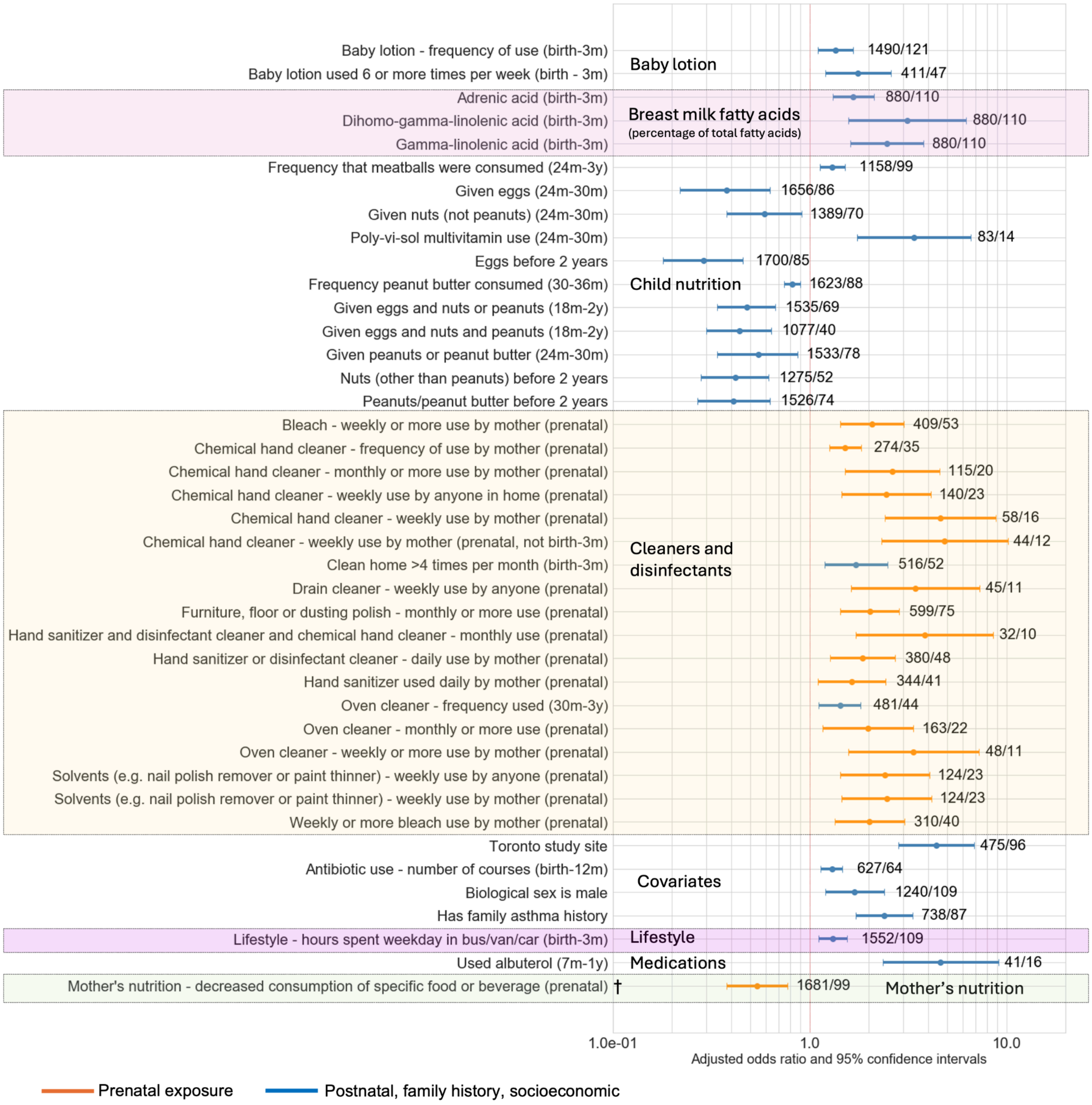


**Supplementary figure 3 | Exposures identified by ExWAS round 2 as being significantly associated with specialist clinician-diagnosed cases of asthma at 3 years of age.** 2343 exposures taking place prior to 3 years of age were assessed for associations with a 3-year asthma diagnosis. Orange and blue horizontal plots are adjusted odds ratios and 95% confidence intervals for prenatal and postnatal exposures, respectively. Numbers to the right of each plot represent total number non-cases and cases subjected to each exposure, respectively. Major groups of exposures are labelled and coloured for additional clarity.

† At enrollment. mother was asked "Since you knew you were pregnant, have you decreased your consumption of any specific foods or beverages?".

### Supplementary figure 4

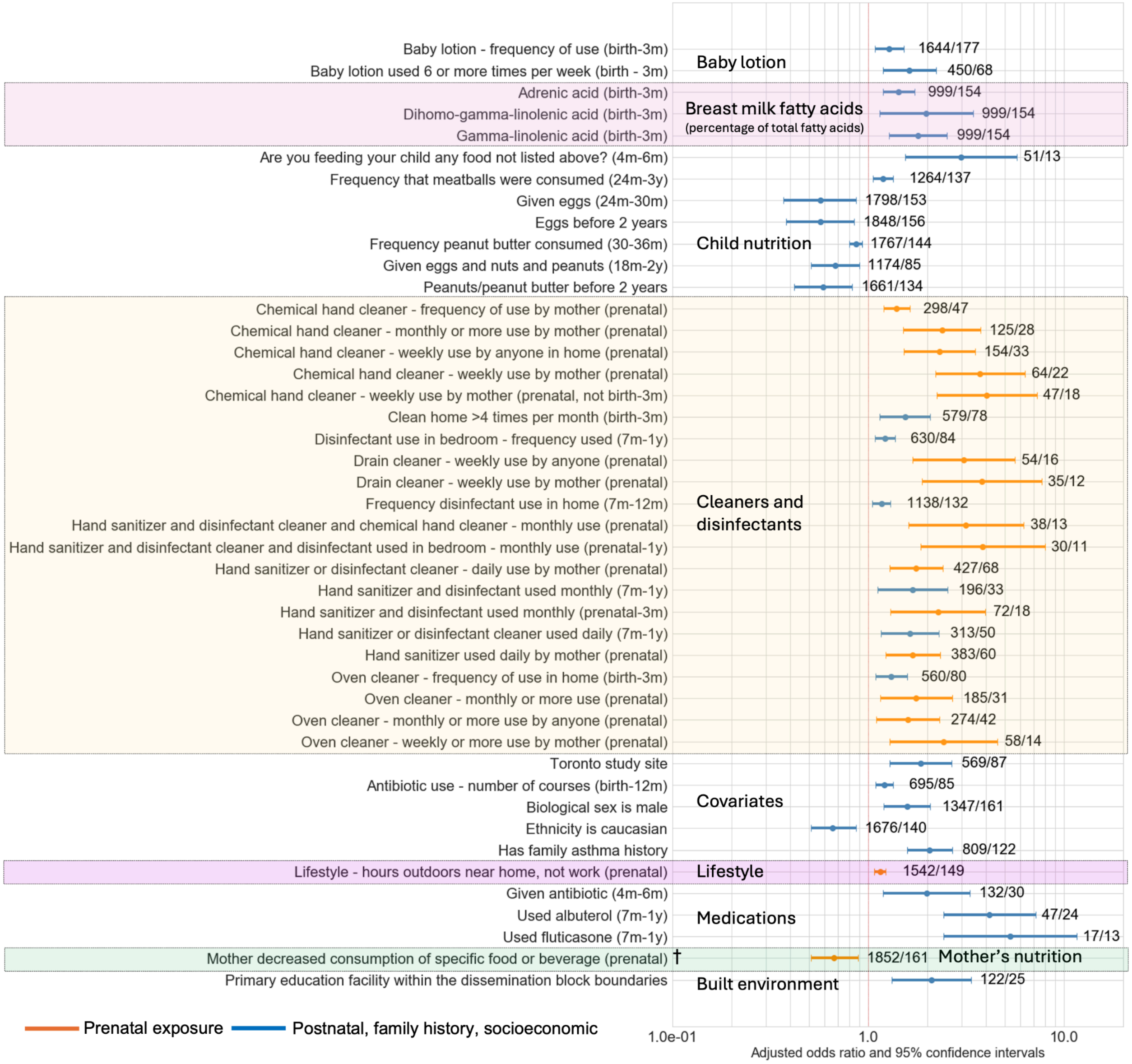


**Supplementary figure 4 | Exposures identified by ExWAS round 2 as being significantly associated with recurrent wheeze at 3 years of age.** 2365 exposures taking place prior to 3 years of age were assessed for associations with 3-year recurrent wheeze. Orange and blue horizontal plots are adjusted odds ratios and 95% confidence intervals for prenatal and postnatal exposures, respectively. Numbers to the right of each plot represent total number non-cases and cases subjected to each exposure, respectively. Major groups of exposures are labelled and coloured for additional clarity.

† At enrollment. mother was asked "Since you knew you were pregnant, have you decreased your consumption of any specific foods or beverages?".

### Supplementary figure 5

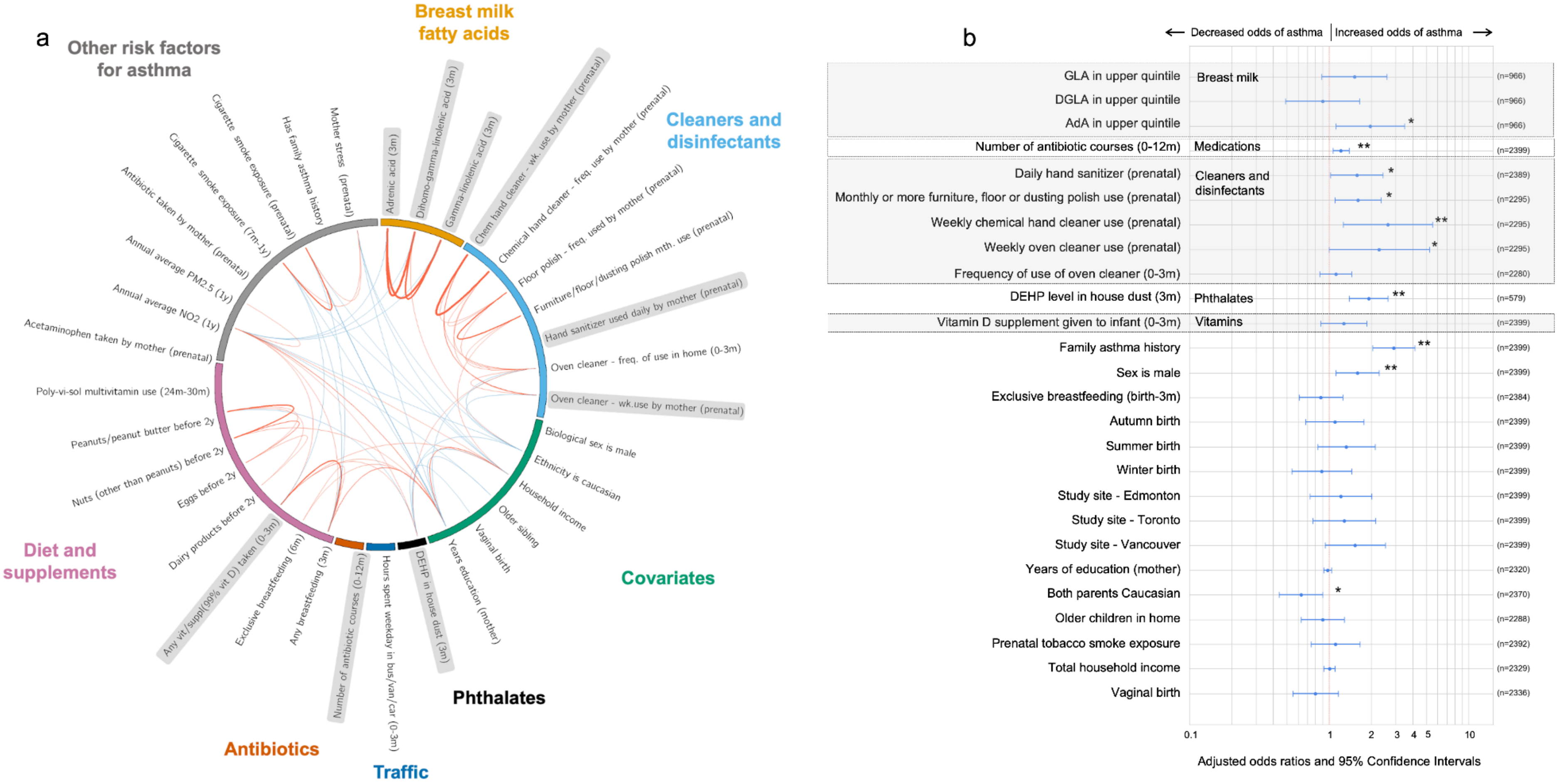

**Supplementary figure 5 | Selection and independent evaluation of prenatal and early-life environmental exposures associated with 5-year asthma. a.** Significant exposures identified by ExWAS, and other risk factors identified in the literature, were grouped into higher level categories (defined by the outer coloured labels), and underwent all versus all correlation analysis against each other and other covariates used in the previous ExWAS analysis. Weaker correlations (positive correlations in red >=0.1; negative correlations in blue <=-0.1) were identified between exposures from different categories while within-category exposures often had correlations with a magnitude greater than 0.2. Line thickness represents the magnitude of the correlation. Exposures that are shaded grey were selected for a confounder analysis and further investigation regarding their association with cytokine, microbiome and epigenetic changes. Abbreviations: Weekly (Wk.), Monthly (Mth.), Frequency (Freq.). **b.** Multivariable logistic regression model assessing the independent associations of multiple prenatal and early-life environmental exposures with 5-year asthma (n=579). Exposures flagged in (a) were entered simultaneously as covariates, along with confounders (mother's education, family income, exclusive breastfeeding status, season of birth, study site, birth mode, family history of asthma, biological sex, older siblings, Caucasian ethnicity, and prenatal tobacco smoke exposure). Separate models were run for each covariate, excluding participants with missing data for that covariate or asthma, while imputing missing values for remaining variables. Numbers to the right indicate the sample size after exclusions. Abbreviations: GLA, gamma linolenic acid; DGLA, dihomo-gamma linolenic acid; AdA, adrenic acid; DEHP, bis(2-ethylhexyl) phthalate; mth, monthly; wk, weekly; NO2, nitrogen dioxide; PM2.5, fine particulate matter in the air, with a diameter of 2.5 micrometers or smaller,. *$p < 0.05$, **$p < 0.01$.

Supplementary figure 6

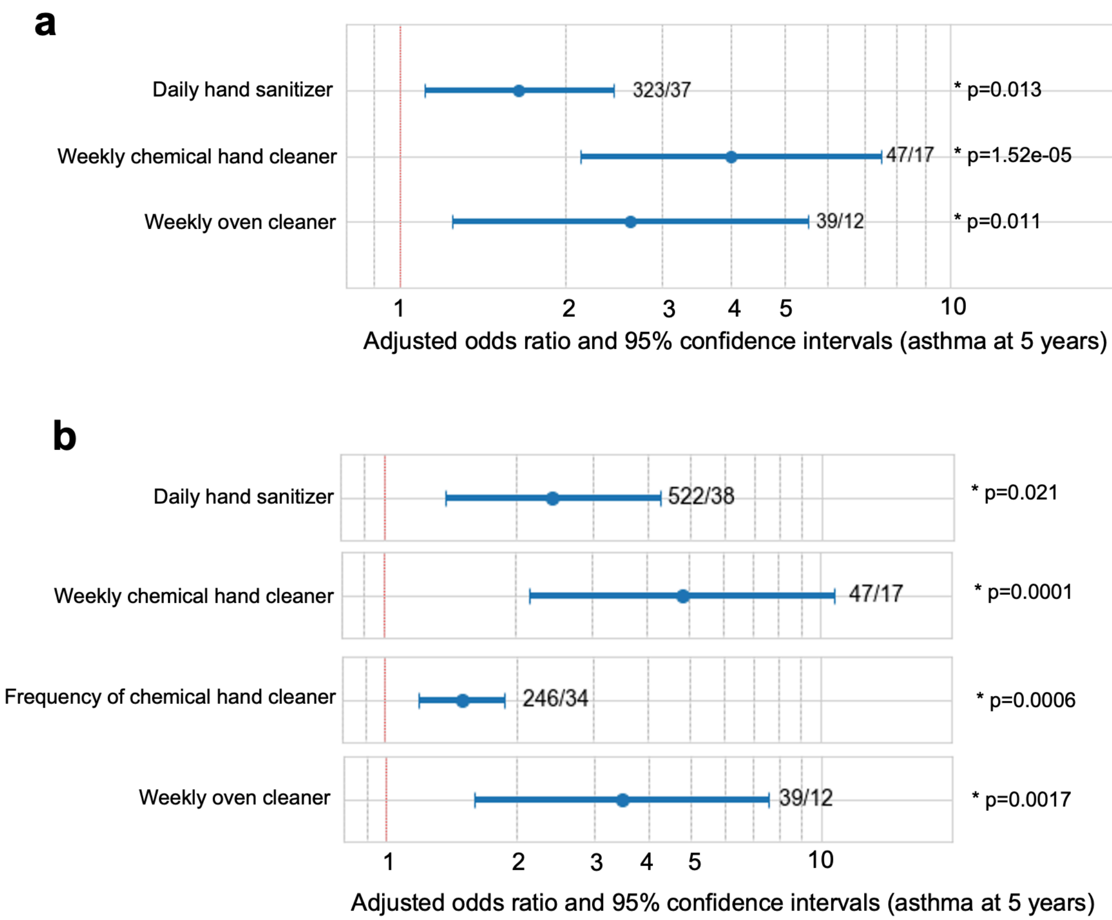

**Supplementary figure 6 | Individual prenatal cleaners and disinfectant use by mother is associated with specialist clinician-diagnosed asthma at 5 years, after adjusting for possible prenatal and postnatal confounders**. **a.** Prenatal cleaner and disinfectant use by mothers (adjusted for postnatal use) is significantly associated with asthma diagnosis at age 5. i. Daily prenatal hand sanitizer use by mother after adjusting for frequency of use reported at 3m, 1y, 3y and 5y. ii. Weekly prenatal chemical hand cleaner use after adjusting for frequency of use reported at 3m and 1y. iii. Frequency of prenatal chemical hand cleaner use after adjusting for frequency of use at 3m and 1y of age. iv.. Weekly prenatal oven cleaner use after adjusting for weekly oven cleaner use reported at 3m, 3y and 5y. Values are adjusted odds ratios and 95% confidence intervals following multivariate logistic regression using definite asthma diagnosis at 5 years as dependent variable. Analysis controlled for postnatal cleaning/disinfectant use and the other covariates used in ExWAS analysis. **b.** Prenatal hand sanitizer, chemical hand cleaner and oven cleaner use by mother, adjusted for the combined effects of all three plus weekly prenatal tile cleaner use that was correlated with them. i. Prenatal hand sanitizer adjusted for weekly prenatal chemical hand cleaner. tile cleaner, and oven cleaner. ii. Prenatal chemical hand cleaner, adjusted for prenatal hand sanitizer, tile cleaner, and oven cleaner. iii. Prenatal oven cleaner, adjusted for prenatal hand sanitizer, tile cleaner, and chemical hand cleaner. Values are adjusted odds ratios and 95% confidence intervals following multivariate logistic regression using definite asthma diagnosis at 5 years as dependent variable.Numbers to the right of each plot represent total number non-cases and cases subjected to each exposure, respectively.

### Supplementary figure 7

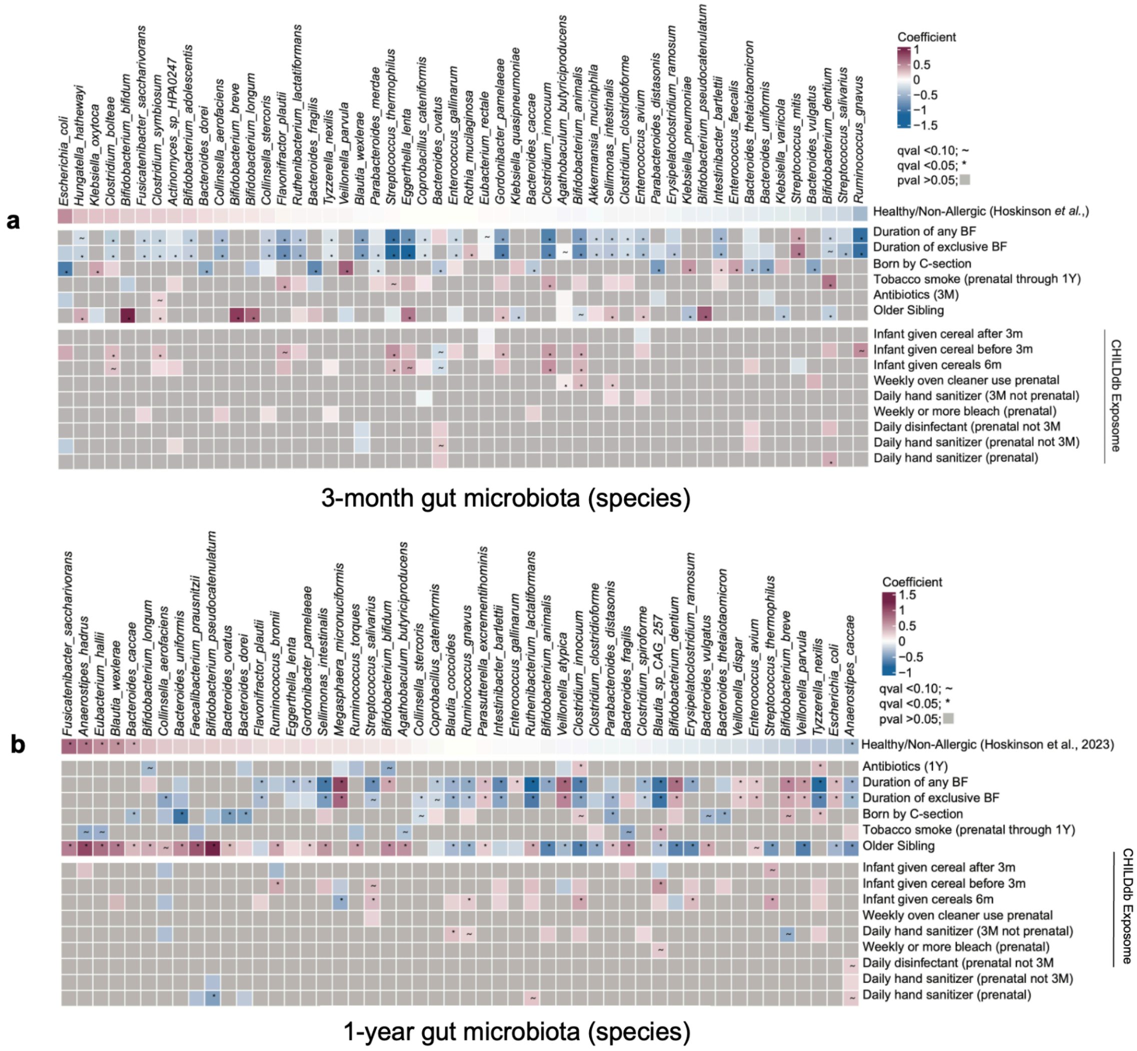


**Supplementary figure 7 | Differentially abundant taxa associated with prenatal cleaner use that were identified in 3-month and 1-year stool samples of CHILD participants.** Taxa identified by Maaslin2 using CLR-transformed ASV counts in linear mixed-effects models examining each of the cleaning product exposures and using study center location as a random effect and adjusting for stool sample of collection age, genetic ancestry of infants PC1-PC10, family income at prenatal enrollment, and biological sex. **a.** Differential abundance in 3-month taxa. **b.** Differential abundance in 1-year taxa.

Supplementary figure 8

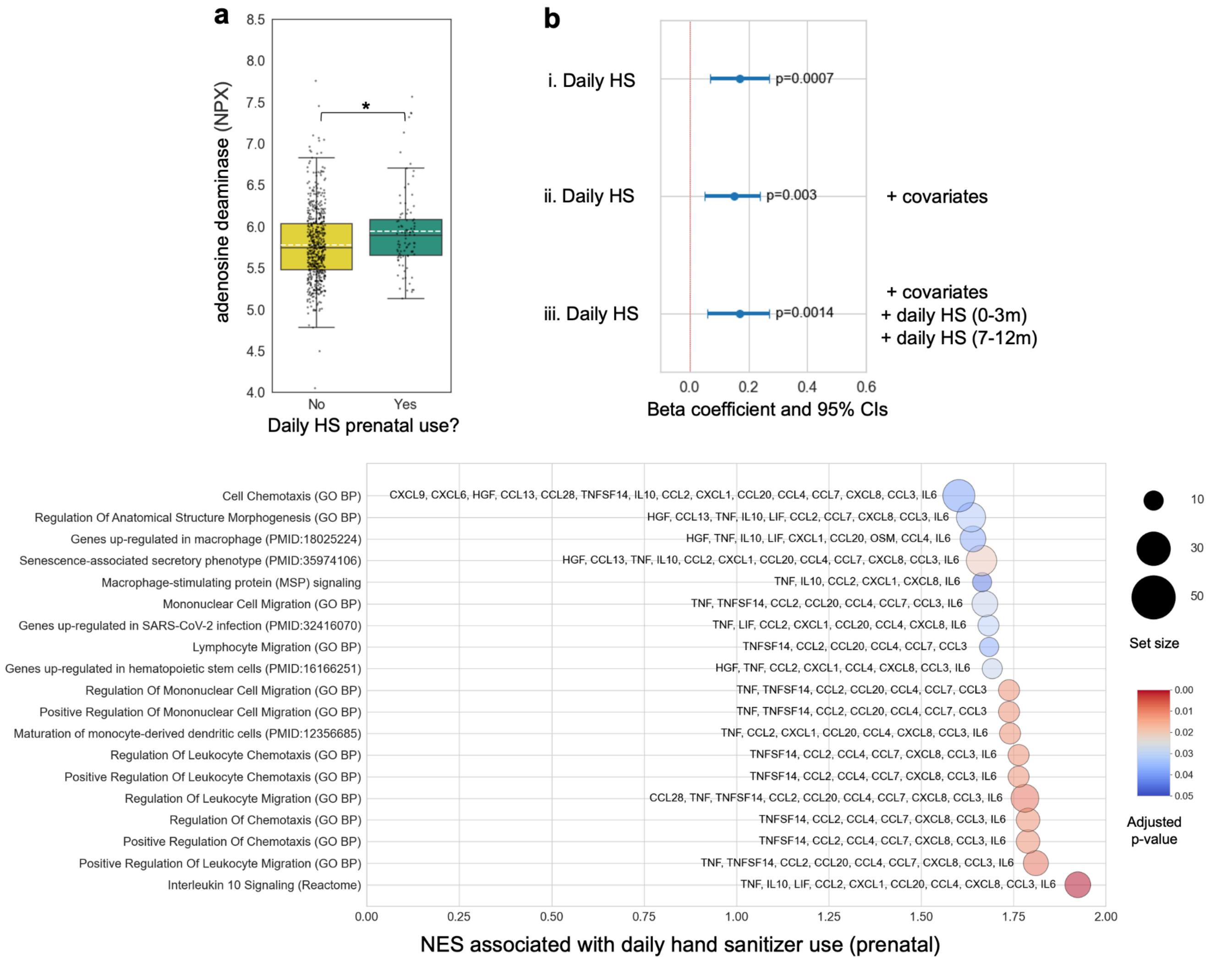

**Supplementary figure 8 | Prenatal daily hand sanitizer associations with cytokines and inflammatory pathways. a.** Children born to mothers reporting prenatal daily use of hand sanitizer had higher adenosine deaminase levels at age 1. Normalized protein expression (NPX) adenosine deaminase levels measured in children's serum collected at 1 year of age, stratified by daily hand sanitizer use (yes/no) by mother during the prenatal period. White dashed line = mean, black solid line = median, * = q<0.05 following correction for multiple testing. **b.** The association between daily prenatal hand sanitizer use and adenosine deaminase remains after adjusting for covariates and daily postnatal use of hand sanitizer. Beta coefficients and 95% confidence intervals for association of prenatal daily hand sanitizer use with adenosine deaminase levels measured in children's serum at 1 year of age. Logistic regression models tested associations between prenatal daily hand sanitizer use and adenosine deaminase using models that were i) unadjusted, ii) adjusted for covariates iii) adjusted for covariates and daily hand sanitizer use (birth to 3 months, 7 months to 1 year). **c.** Gene Set Enrichment Analysis (GSEA) of 5-year serum identifies prenatal daily hand sanitizer use associated with enriched Interleukin-10 signalling, maturation of monocyte-derived dendritic cells, and regulation of chemotaxis (adjusted p-value <0.05).

#### Supplementary table 3

**Supplementary table 3 | Evidence for core enrichment biomarker associations with Th2-high, Th2-low, Th2-low(Th1-high subset) immune profiles and airway remodeling.** Core enrichment biomarkers (i.e. leading edge genes) that contributed most to enrichment of biological pathways from MSigDB database annotations were curated for additional evidence in the scientific literature that associated them with common immune profiles associated with asthma and airway remodeling.

| Cytokine | Synonym | T2-high profile | T2-low profile | T2-low (T1-high) profile | Airway Remodeling |
|---|---|---|---|---|---|
| CCL11 | Eotaxin-1 | 173 | | | |
| CCL19 | MIP-3β | 174 | | | |
| CCL2 | MCP-1 | 175 | 176 177 178 | | |
| CCL20 | MIP-3α | | 179,180 | | |
| CCL25 | TECK | 28 | | | |
| CCL3 | MIP-1α | | 176 82,181 | | |
| CCL4 | MIP-1β | | 176 182 | | |
| CCL7 | MCP-3 | 183 | 178,184–186 | | |
| CCL8 | MCP-2 | | 176,187,188 | | |
| CX3CL1 | Fractalkine | 189,190 | | | |
| CXCL1 | GROα, NAP-3 | | 177,181,191 | | |
| CXCL10 | IP-10 | | 176,184 | 84,116,192 | |
| CXCL11 | I-TAC, IP-9 | | | 84,116,192 | |
| CXCL5 | ENA78 | 117,193 | 118,194 | | |
| CXCL6 | GCP-2 | | 195,196 | | |
| CXCL8 | Interleukin-8 | | 82,176,191,197,198 | | |
| CXCL9 | MIG | | | 84,116 | |
| GDNF | | | | | 199,200 |
| IFNG | IFN-γ | | 82 | 116 | |
| IL13 | Interleukin-13 | 201,202 | 203 | | |
| IL6 | Interleukin-6 | | 82,176,177,198 | | |
| TNF | TNF-alpha | | 82,176 | | |
| TNFSF11 | | | | | 204 |
| TNFSF14 | LIGHT | | | | 205,206 |
| VEGFA | VEGF-A | 207,208 | | | 208–210 |

Supplementary figure 9

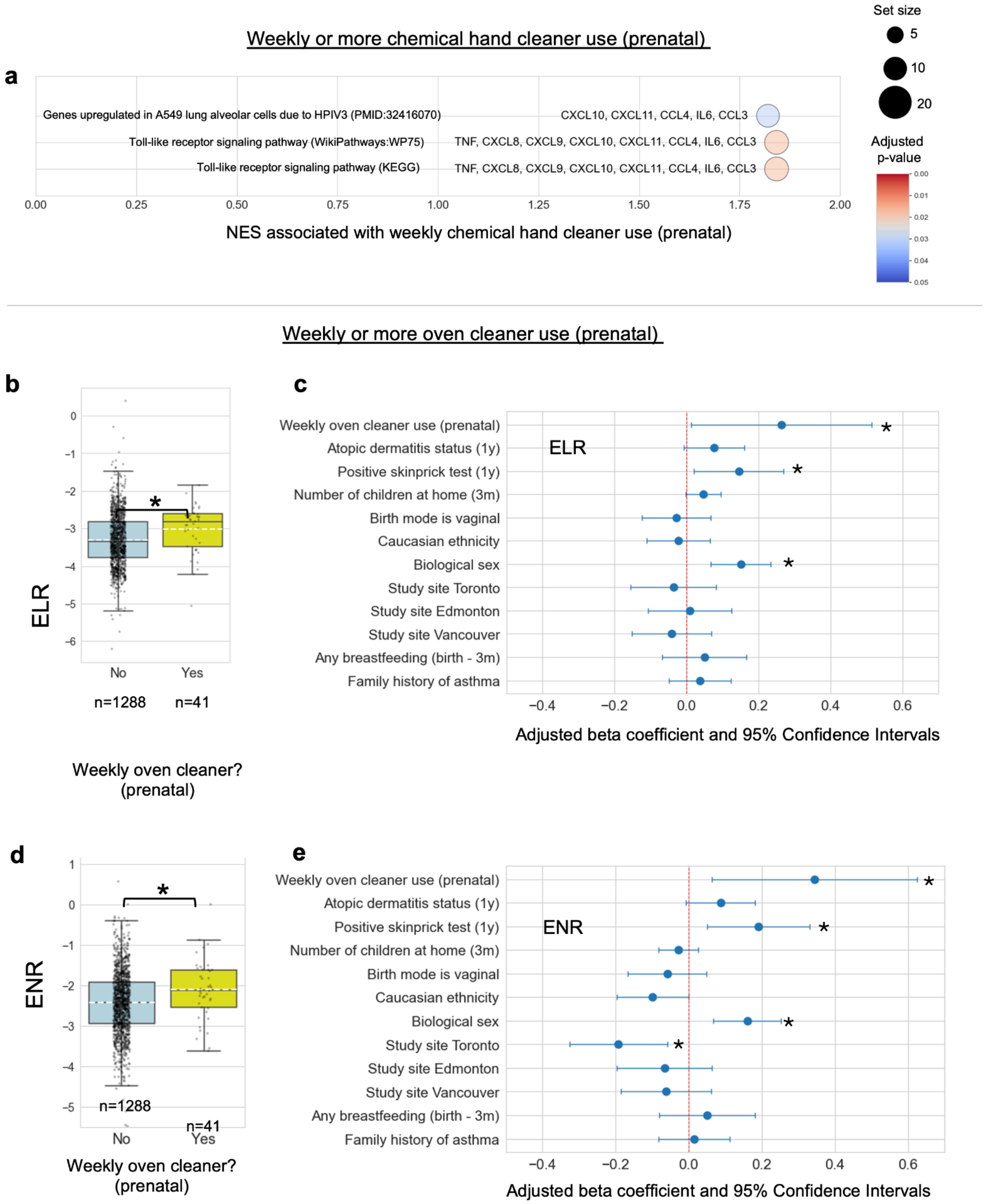

**Supplementary figure 9 | Associations between prenatal use of cleaning products (chemical hand cleaner, oven cleaner) by mother and inflammatory pathways/markers. a.** Gene set enrichment analysis (GSEA) of 1-year serum cytokines identified association between prenatal weekly chemical hand cleaner use by mother and inflammatory pathways including the toll-like receptor signalling pathway (all with adjusted p-value <0.05). **b.** Mean log eosinophil/lymphocyte ratios (ELR) at age 1 year in children of mothers who used oven cleaner on a weekly or more basis during the prenatal period (yellow) is higher than those that did not (blue). *Statistically significant after FDR correction at $q < 0.05$. **c.** Prenatal weekly oven cleaner use by mother is associated with 1-year ELR after adjusting for atopic dermatitis status at 1 year, any positive skin prick test for food and aeroallergens, number of children at home, birth mode, biological sex, study site, any breastfeeding at 3 months, and family asthma history. *$p < 0.05$. **d.** Mean log eosinophil/neutrophil ratios (ENR) in children of mothers who used oven cleaner on a weekly or more basis during the prenatal period (yellow) are higher than those that did not (blue). *Statistically significant after FDR correction at $q < 0.05$. **e.** Prenatal weekly oven cleaner use by mother is associated with 1-year ENR after adjusting for atopic dermatitis status at 1 year, any positive skin prick test for food and aeroallergens, number of children at home, birth mode, biological sex, study site, any breastfeeding at 3 months, and family asthma history. *$p< 0.05$. Abbreviations: eosinophil/lymphocyte ratio (ELR), eosinophil/neutrophil ratio (ENR)

### Supplementary figure 10

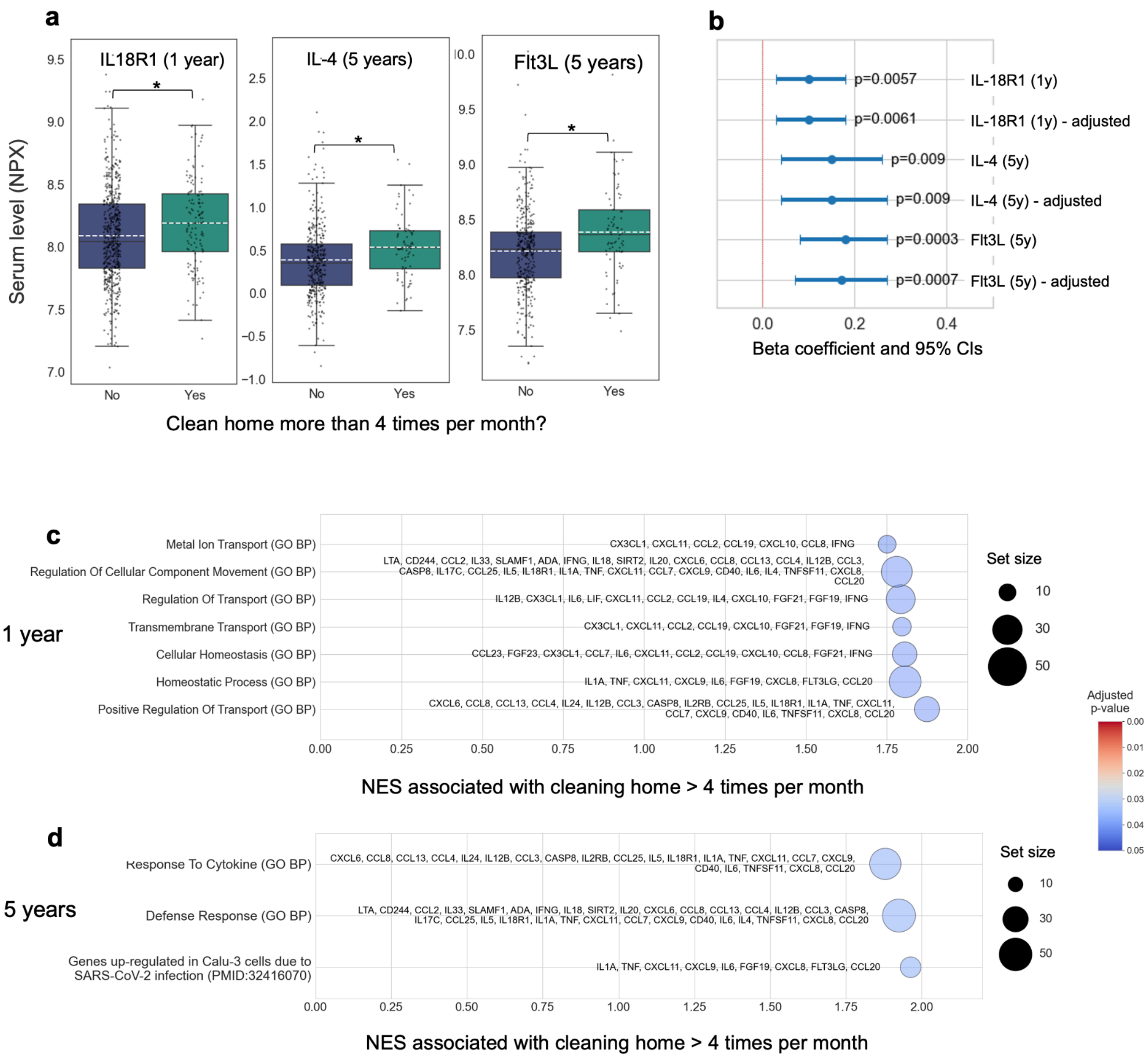

**Supplementary figure 10 | General home cleaning >=4 times per month (between birth and 3 months of age) is associated with increased serum cytokine levels and enriched biological pathways.** **a**. Children's serum IL-18R1 (1-year), IL-4 (5-year). and Flt3L (5-year) are higher in children from homes where cleaning took place more than 4 times per month between birth and 3 months of age. * Significant by Mann-Whitney U test and corrected for multiple testing using Benjamini Hochberg ($q<0.05$) and validated using ANCOVA controlling for covariates. **b.** After controlling for covariates, children's serum IL-18R1 (1-year), IL-4 (5-year). and Flt3L (5-year) remain significantly higher in children from homes where cleaning took place more than 4 times per month. Beta coefficients and 95% confidence intervals are shown for unadjusted and adjusted multiple linear regression models. Covariates used for multiple linear regression were study site, biological sex, ethnicity, BMI at year of serum collection and breastfeeding status (any breastfeeding) at 3 months of age. **c.** From 1-year serum cytokine levels, gene set enrichment analysis (GSEA) identified associations between cleaning homes more than 4 times per month and regulation of transport and cellular homeostasis (all with adjusted p-value <0.05). **d.** From 5-year serum cytokine levels, gene set enrichment analysis (GSEA) identified associations between cleaning homes more than 4 times per month and the defense response and response to cytokine pathways (all with adjusted p-value <0.05).

#### Supplementary table 4

**Supplementary table 4 | Weekly prenatal oven cleaner use by mother is correlated with prenatal chemical hand cleaner and tile cleaner use, and postnatal oven cleaner use.** Spearman rank correlation coefficients (ρ) for analysis comparing weekly prenatal oven cleaner use by mother versus the entire 2954-variable ExWAS dataset. False discovery rate was controlled for using the Benjamini-Hochberg method and significant correlations with an absolute value for Spearman ρ >=0.1 were retained.

| Correlated variable | n | Correlation coefficient | p-value | q |
|---|---|---|---|---|
| Weekly or more oven cleaner use by mother (birth-3m) | 2874 | 0.279 | 1.43716E-52 | 4.31E-49 |
| Weekly chemical hand cleaner use by mother (prenatal) | 3062 | 0.246 | 1.90563E-43 | 2.86E-40 |
| Weekly or more oven cleaner use (30m-3y) | 3060 | 0.217 | 5.70797E-34 | 5.71E-31 |
| Monthly or more chemical hand cleaner use by mother (prenatal) | 3062 | 0.209 | 1.48024E-31 | 9.38E-29 |
| Chemical hand cleaner - weekly use by mother (prenatal, not birth-3m) | 3062 | 0.209 | 1.56354E-31 | 9.38E-29 |
| Weekly tile cleaner use (prenatal) | 3062 | 0.203 | 5.62388E-30 | 2.81E-27 |
| Hand sanitizer AND disinfectant cleaner AND chemical hand cleaner - monthly use (prenatal) | 3062 | 0.199 | 7.77778E-29 | 3.33E-26 |
| Weekly chemical hand cleaner use by anyone in home (prenatal) | 3062 | 0.191 | 1.92621E-26 | 7.22E-24 |
| Choline in mother's serum (prenatal) | 296 | 0.172 | 0.0030639 | 0.045488 |
| Frequency of use of oven cleaner in home (30m-3y) | 2231 | 0.171 | 3.75631E-16 | 8.67E-14 |
| Monthly or more oven cleaner use (birth-3m) | 2874 | 0.169 | 7.21936E-20 | 2.41E-17 |
| Frequency of chemical hand cleaner use by mother (prenatal) | 3054 | 0.156 | 3.46235E-18 | 1.04E-15 |
| Weekly chemical hand cleaner use by mother (prenatal, not 6m-12m) | 3062 | 0.149 | 1.14982E-16 | 3E-14 |
| Monthly or more brass polish use (prenatal) | 3062 | 0.149 | 1.20105E-16 | 3E-14 |
| Frequency of use of oven cleaner in home (birth-3m) | 2863 | 0.146 | 3.82965E-15 | 8.2E-13 |
| Feeding method is bottle with soft plastic bottle liner (18m-2y) | 2222 | 0.139 | 5.19832E-11 | 7.79E-09 |
| Weekly drain cleaner use by anyone (prenatal) | 3062 | 0.138 | 1.55461E-14 | 3.11E-12 |
| Child given soy milk (4y-5y) | 466 | 0.138 | 0.002900598 | 0.043713 |
| Weekly chemical hand cleaner use by mother (6m-12m) | 2493 | 0.137 | 6.74515E-12 | 1.06E-09 |
| Weekly or more toilet bowl cleaner use (prenatal) | 3062 | 0.130 | 5.68112E-13 | 1.06E-10 |
| Frequency of use of chemical hand cleaner (7m-1y) | 2493 | 0.127 | 1.83741E-10 | 2.5E-08 |
| Weekly or more bleach use by mother (prenatal) | 3062 | 0.127 | 1.75875E-12 | 3.1E-10 |
| Frequency of use of bathroom tile cleaner (prenatal) | 3056 | 0.125 | 4.22844E-12 | 7.05E-10 |
| Weekly tile cleaner use (birth-3m) | 2874 | 0.117 | 2.73062E-10 | 3.41E-08 |
| Frequency floor cleaner used by anyone (prenatal) | 3054 | 0.116 | 1.09369E-10 | 1.56E-08 |
| Weekly or furniture, floor or dusting polish more use (prenatal) | 3062 | 0.113 | 4.13889E-10 | 4.97E-08 |
| Air fresheners - liquid or solid - frequency of use (birth-3m) | 2866 | 0.111 | 2.87744E-09 | 3.2E-07 |
| Frequency of use of floor cleaners by anyone in home (prenatal) | 3053 | 0.110 | 1.23108E-09 | 1.42E-07 |
| Feeding method is bottle with soft plastic bottle liner (2y-30m) | 2114 | 0.109 | 4.62088E-07 | 2.89E-05 |
| Weekly chemical hand cleaner use by mother (birth-3m) | 2874 | 0.109 | 5.19142E-09 | 5.37E-07 |
| Weekly chemical hand cleaner use (birth-3m) | 2874 | 0.109 | 5.19142E-09 | 5.37E-07 |
| Frequency of use of gas space heater (birth-3m) | 2861 | 0.108 | 6.22166E-09 | 6.22E-07 |
| Glass/ceramic used for food storage (2y-30m) | 2117 | -0.100 | 3.716E-06 | 0.000183 |
| Ethnicity is caucasian | 3040 | -0.115 | 2.089E-10 | 2.72E-08 |
| Meals per week consuming foods purchased from grocery store (24m-3y) | 2266 | -0.117 | 2.49294E-08 | 2.34E-06 |
| Child nutrition - grains introduced before 2 years | 2230 | -0.122 | 6.99148E-09 | 6.76E-07 |
| Arginine in mother's serum (prenatal) | 296 | -0.171 | 0.003180293 | 0.046525 |

#### Supplementary figure 11

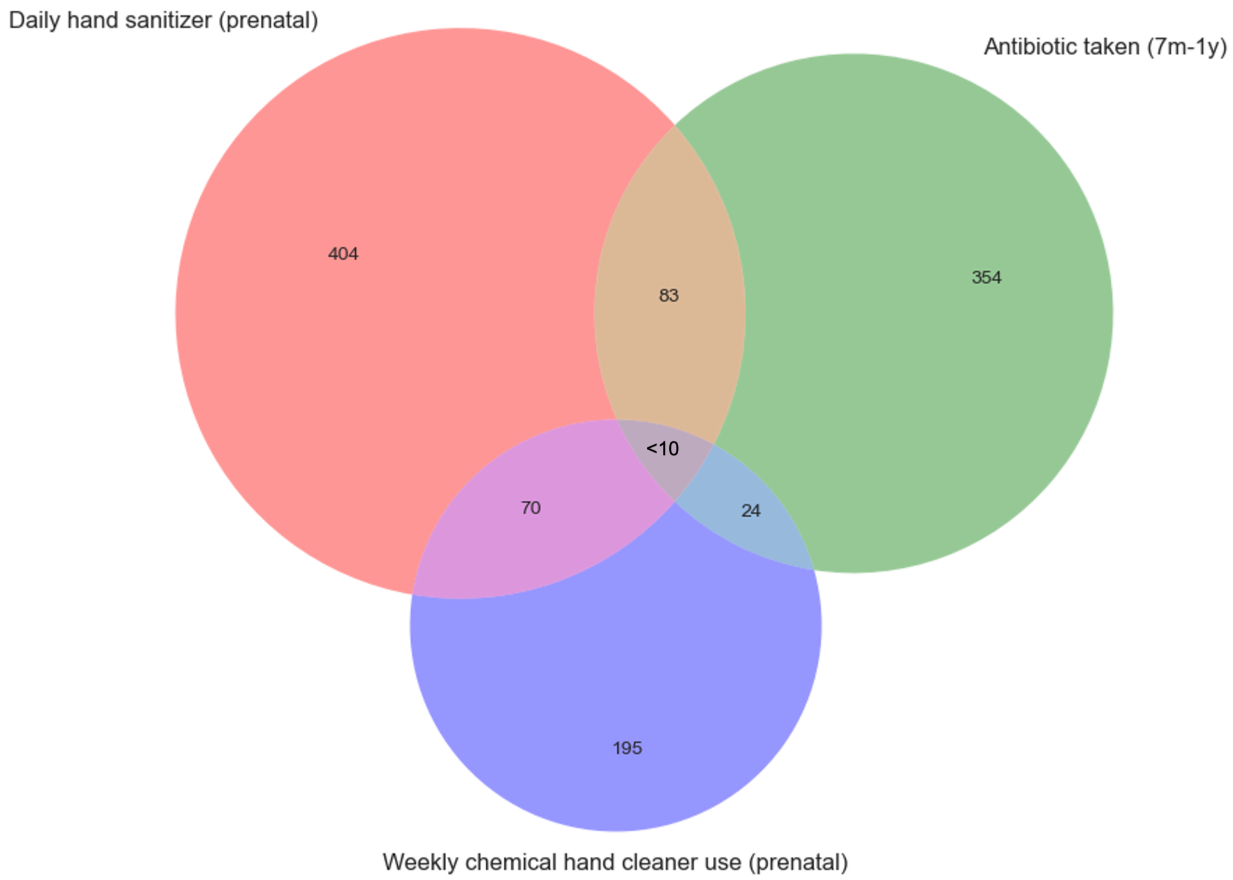


**Supplementary figure 11 | Limited overlap in weekly or more chemical hand cleaner use (prenatal), daily hand sanitizer use (prenatal), and infants being given at least one antibiotic (between 7 months and 1 year of age).** A Venn diagram was constructed to demonstrate the degree of overlap in three exposures associated with development of asthma at 5 years of age.

#### Supplementary figure 12

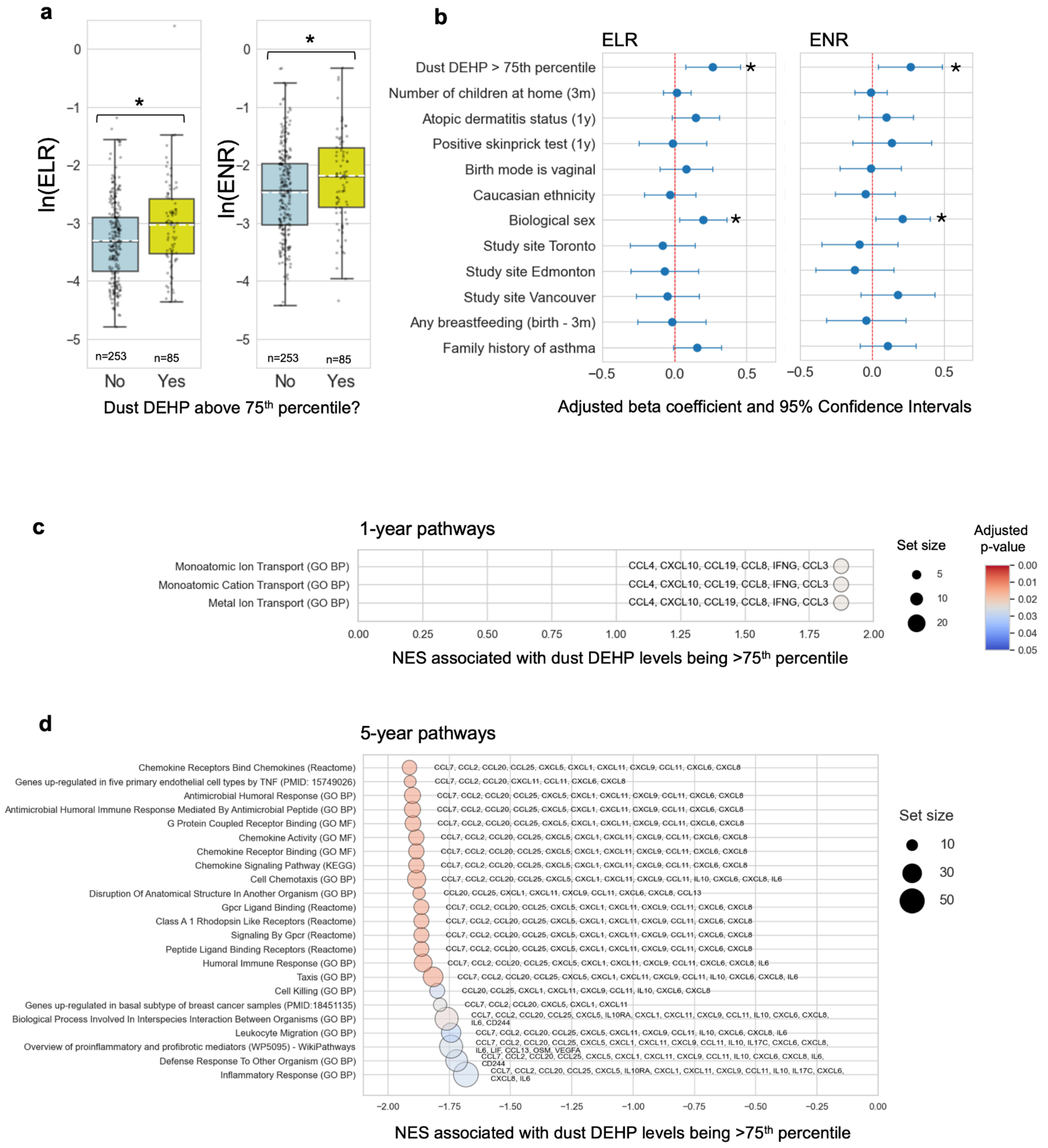

**Supplementary figure 12 | DEHP levels in house dust are associated with increased ELR and ENR at 1 year, positively-enriched ion transport at 3 months, and negative enrichment of chemotaxis/cell migration pathways at 5 years. a**. Mean natural log-transformed ELR and natural log-transformed ENR in children exposed to dust DEHP levels not above the 75th percentile (blue) versus dust DEHP levels above the 75th percentile (yellow). *Statistically significant after FDR correction ($q < 0.05$). **b.** Association between ELR or ENR at age 1 and DEHP in the upper quartile, adjusted for number of children in home, atopic dermatitis status at 1 year, positive skin prick test for at least one food or aeroallergen, birth mode, caucasian ethnicity, biological sex, study site, any breastfeeding at 3 months, and family history of asthma (*$p<0.05$). **c.** Gene set enrichment analysis (GSEA) for house dust DEHP levels in upper quartile versus those that were not reveals pathways enriched from biomarkers increased in 1-year serum of participants in the former group. **d.** GSEA for house dust DEHP levels in upper quartile versus those that were not reveals pathways enriched from biomarkers decreased in 5-year serum of participants in the former group. Negative NES indicate downregulation of the pathway in exposed individuals, while positive NES scores indicate upregulation of the pathway in exposed individuals. Set size represents the number of proteins in the specific molecular signatures database gene set after filtering out genes not included in the serum cytokine dataset. Core enrichment genes that contribute most to the enrichment signal are listed to the side of each bubble. Abbreviations: bis(2-ethylhexyl) phthalate (DEHP), eosinophil/lymphocyte ratio (ELR), eosinophil/neutrophil ratio (ENR), normalized enrichment scores (NES).

#### Supplementary figure 13

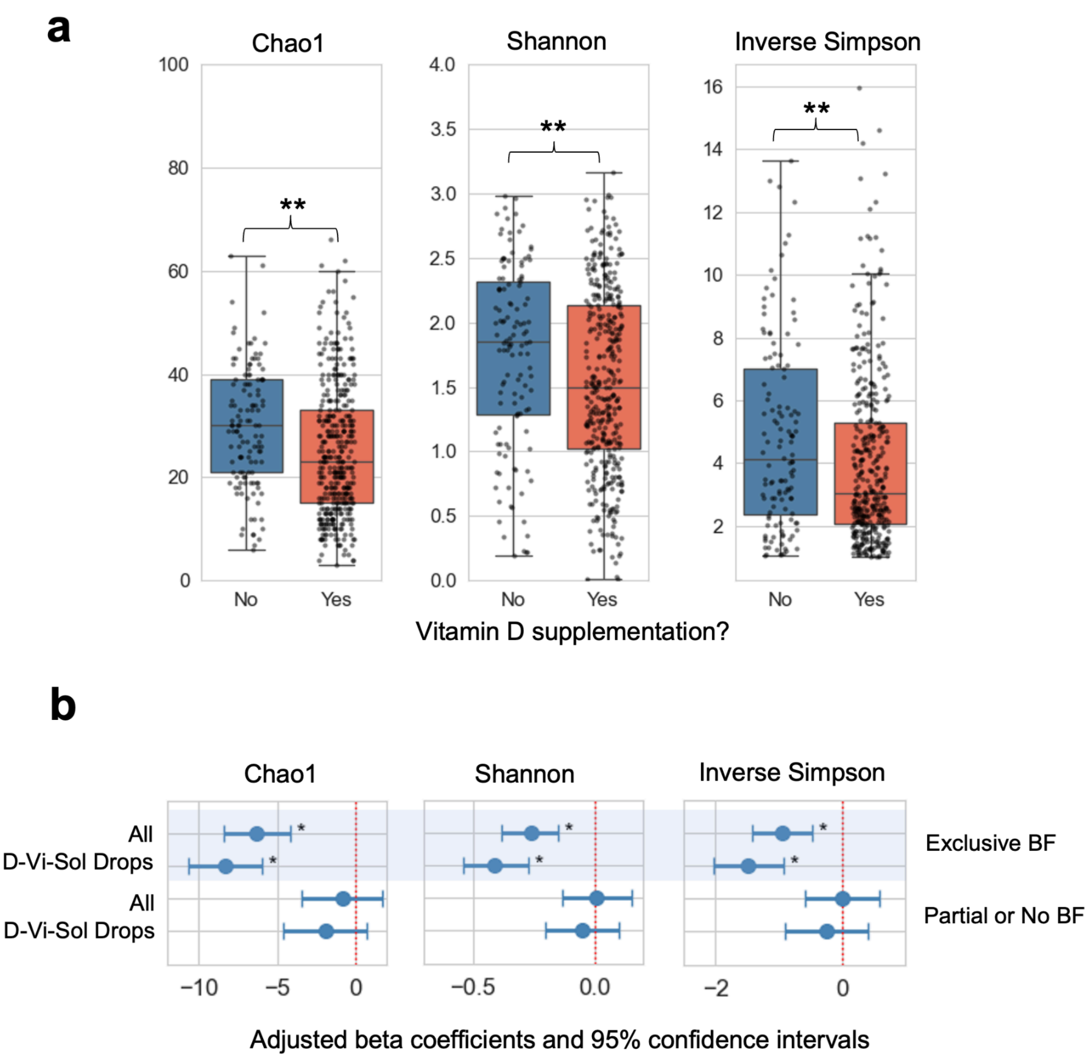


**Supplementary figure 13 | Infants given vitamin D between birth and 3 months of age exhibit gut microbiome decreased diversity**. **a.** Alpha diversity (Chao1, Shannon, and Inverse Simpson) for gut microbiota in exclusively-breastfed infants at age 3 months that were or were not given vitamin D supplements between birth and 3 months (**p <0.01, Mann-Whitney U test). **b.** Linear mixed effects modelling used to infer associations between vitamin D supplementation (0-3m) and alpha diversity of 3-month gut microbiota in infants stratified by being exclusively breastfed (BF) or not exclusively breastfed at the age of 3 months, after controlling for confounders and correcting for FDR using the Benjamini-Hochberg method (*q-val <0.05).

#### Supplementary figure 14

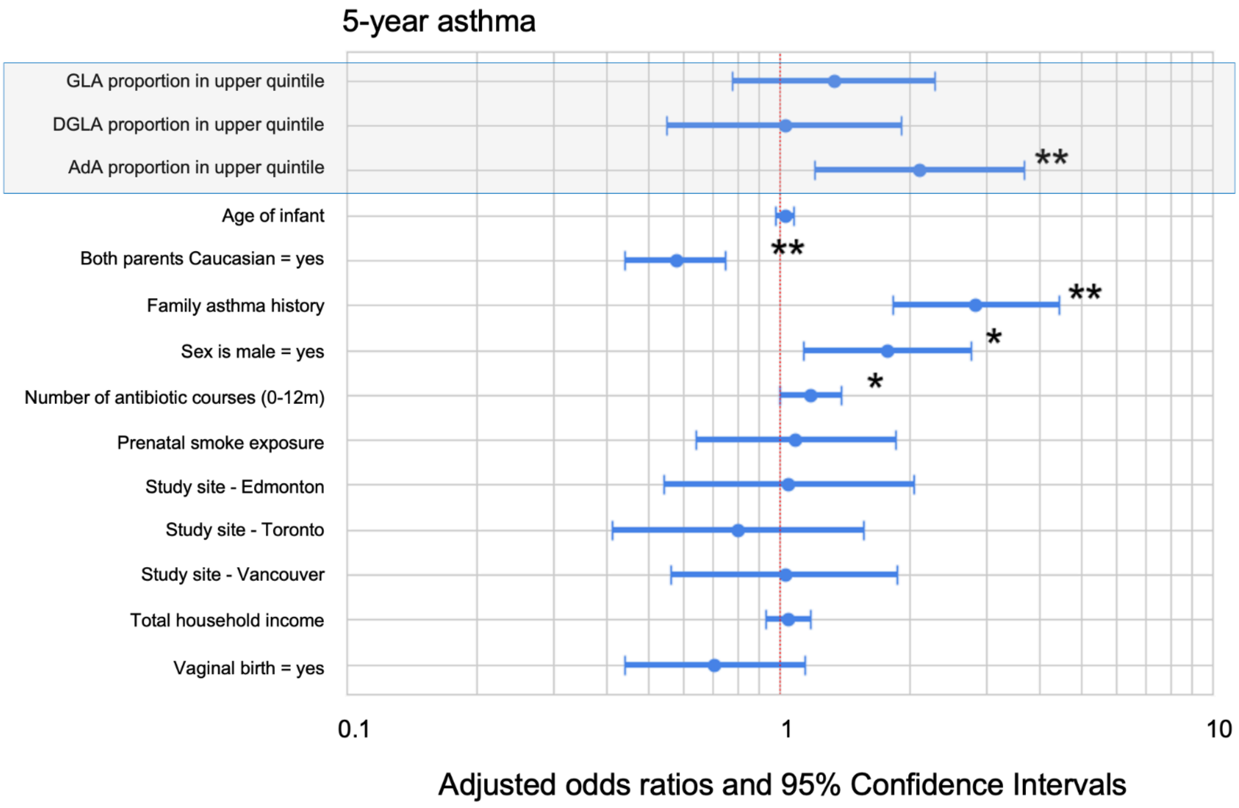


**Supplementary figure 14 | High levels of human milk AdA are positively associated with specialist clinician-diagnosed asthma at 5 years (excluding children breastfed less than 3 months). a.** Forest plot of associations between GLA, DGLA, and AdA concentrations being in the upper quintile (in grey shaded box) and 5 -year asthma after mutually adjusting for levels of all 3 fatty acids and expanded covariates including infant age at time of milk sample collection (n=911 children). Abbreviations: gamma linolenic acid (GLA), dihomo-gamma linolenic acid (DGLA), adrenic acid (AdA). P-values: * p <0.05, ** p < 0.01.

Supplementary figure 15

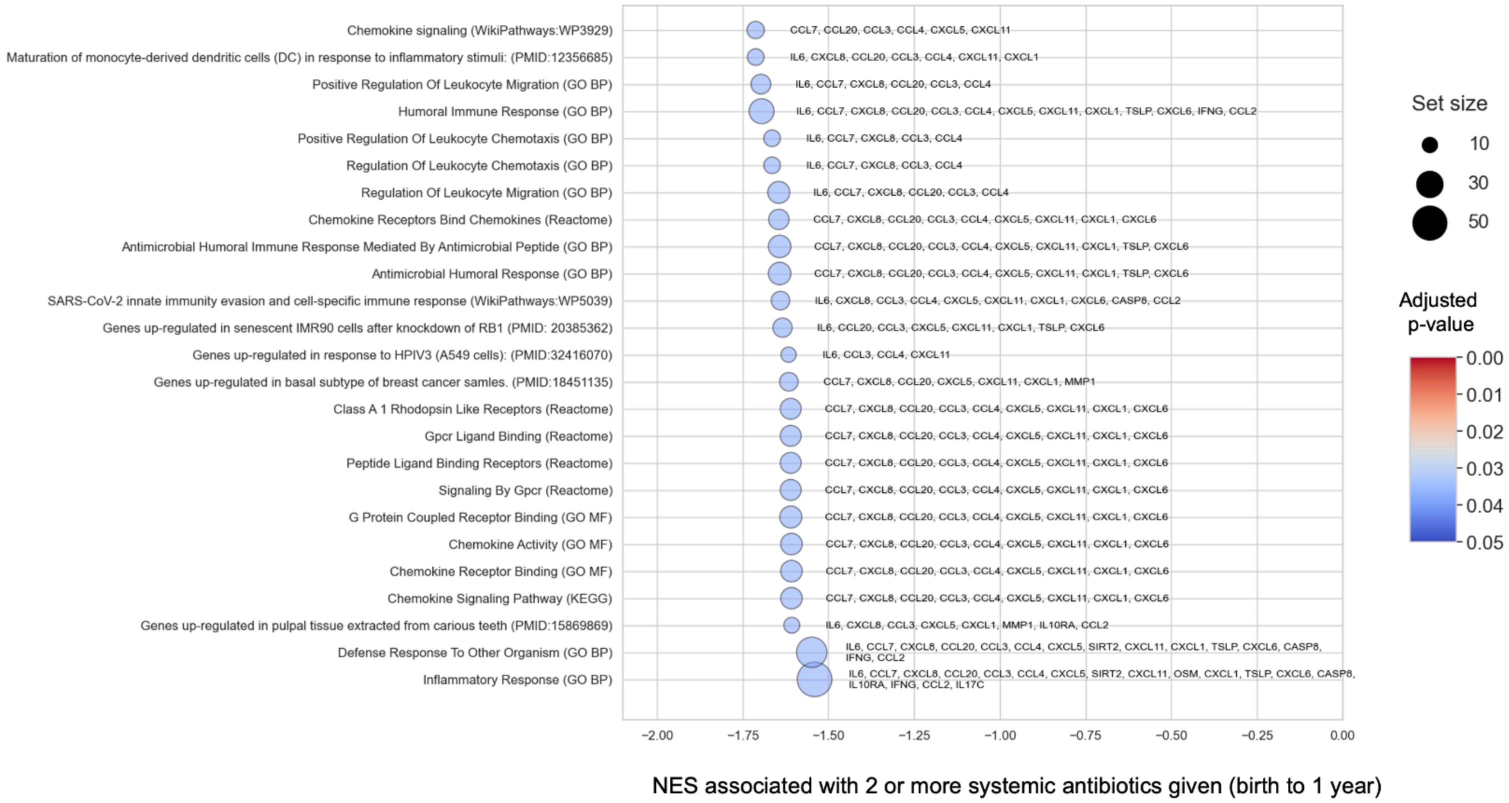

**Supplementary figure 15 | Gene set enrichment analysis (GSEA) of biological pathways associated with the children's 5-year serum cytokine dataset in response to the infant being given two or more systemic antibiotics between birth and 1 year of age**. Top significantly-enriched pathways for infants receiving 2 courses of systemic antibiotics between birth and 1 year of age (n=20) relative to those that received zero or one course (n=412). Negative normalized enrichment scores (NES) indicate pathways enriched from biomarkers that were downregulated in 5-year serum samples from participants given two courses of systemic antibiotics. Diameter of circles indicates set size that reflects the number of proteins in the specific molecular signatures database gene set after filtering out genes not included in the serum cytokine dataset. Core enrichment genes that contribute most to the enrichment signal are listed to the side of each bubble.

#### Supplementary figure 16

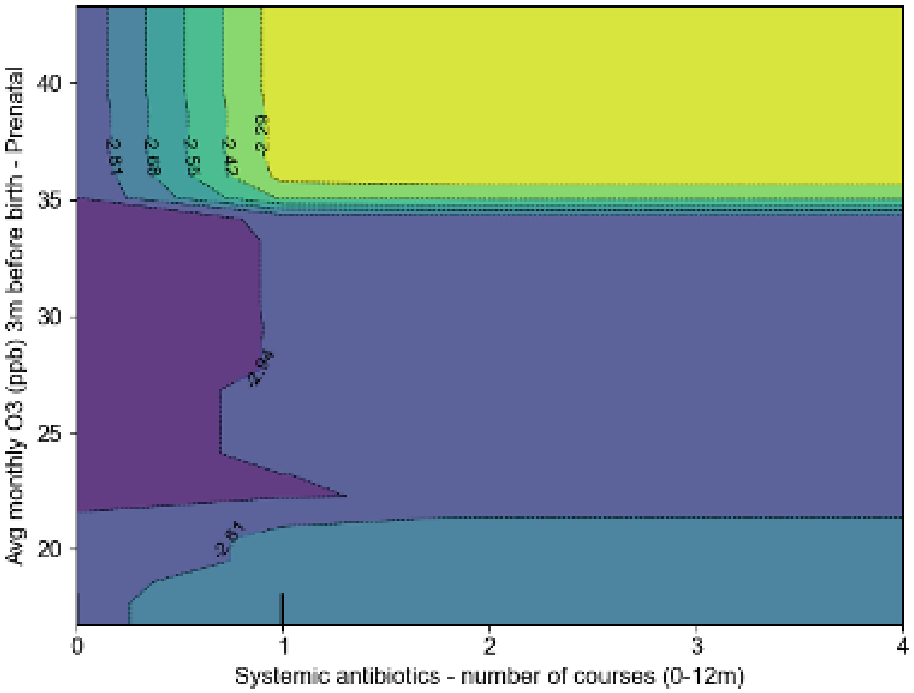


**Supplementary figure 16 | Two-dimensional partial dependence plot for interaction between average monthly O3 (ppb) levels measured 3 months prior to birth and number of courses of systemic antibiotics taken by the child between birth and 1 year of age.** Colormap represents predicted values on the log-odds scale, where higher values (yellow) indicate increased odds of asthma and lower values (purple) indicate decreased odds.
